# A general computational framework for COVID-19 modelling with applications to testing varied interventions in education environments

**DOI:** 10.1101/2021.03.08.21253122

**Authors:** Joshua W. Moore, Zechariah Lau, Katerina Kaouri, Trevor C. Dale, Thomas E. Woolley

## Abstract

We construct a spatially-compartmental individual-based model of the spread of Covid-19 in indoor spaces. The model can be used to predict the infection rates in a variety of locations when various non-pharmaceutical interventions (NPIs) are introduced. Tasked by the Welsh Government, we apply the model to secondary schools and Further and Higher Education environments. Specifically, we consider student populations mixing in a classroom and in halls of residence. We focus on assessing the potential efficacy of Lateral Flow Devices (LFDs) when used in broad-based screens for asymptomatic infection or in ‘test-to-release’ scenarios in which individuals who have been exposed to infection are released from isolation given a negative result. LFDs are also compared to other NPIs; we find that, although LFD testing can be used to mitigate the spread of Covid-19, it is more effective to invest in personal protective equipment, e.g. masks, and in increasing ventilation quality. In addition, we provide an open-access and user-friendly online applet that simulates the individual-based model, complete with user tutorials to encourage the use of the model to aid educational policy decisions as input infection data evolves (https://bit.ly/CV19_INTER_IBM).

## 1 Introduction

When the Covid-19 pandemic started sweeping across the globe [1, 2, 3], governments tried to stem spread by imposing national lockdowns during which essential sectors, such as education, were shut down, or moved online [4]. Following successive epidemic waves there have been extensive efforts to get students, across all age ranges, back into schools, colleges and universities [5, 6, 7, 8]. Primarily, the arguments for reopening education environments have focused on the benefits of education for the development of skills and knowledge as well as for the maintenance of good mental health [9, 10, 11, 12].

There has been a vigorous debate about the role of schools and universities in disease transmission and, as such, a range of non-pharmaceutical interventions (NPIs) have been proposed and implemented in the education sector [13, 14, 15]. In the UK, the focus has been on using Lateral Flow Devices (LFDs) as a means of detecting infected individuals to better manage infection [16, 17, 18]. It has been suggested that a negative result from an LFD might be used to discontinue the self-isolation of people who are a close contact of an infected individual [19, 20, 21], with trepidation caused by the fact that LFDs have a high false negative rate [22]. We, thus, investigated how beneficial LFDs can be in comparison to other NPIs, such as wearing masks or improving ventilation, particularly as recent literature has reported that despite the success of the vaccination programme which led to a reduction in compliance to contact and isolation policies [23]. Moreover, vaccinated individuals can still contract and transmit the infection, specifically if the individual has underlying health conditions and/or is taking medications that impair the immune system [24, 25]. The Centers for Disease Control and Prevention state such an individual should “should continue to take all precautions recommended for unvaccinated people, including wearing a well-fitted mask, until advised otherwise by their healthcare provider”[26]. Hence there is need for continuing to study NPIs to identify their optimal use, with emphasis on their roles in educational environments.

There has been recent efforts to physically quantify the use of LFDs in a ‘test-to-release’ strategy by means of a national experiment of 204 UK secondary schools over a six week period [27]. The study compared the ‘test-to-test’ strategy against the existing alternative of close-contact isolation, measuring the number of infections confirmed by a positive PCR test within each of the schools. The authors conclude that the use of LFDs in a ‘test-to-release’ isolation policy did not significantly increase the number infections within the selected cohorts and therefore suggested its viability to reduce absences from education. However, the study was conducted when the disease was at its lowest prevalence and least transmissive in England in 2021 [28, 29] and therefore does not indicate whether the results of the study are viable in a highly infectious environment, particularly with the estimated false-negative rate of self-administered LFDs [22]. In addition, the number of infections in both the control and intervention groups where scaled against the local community cases, yet due to the high asymptomatic rate of cases for people aged 12-18 [30], the effects of greater prevalence would not be observed within the schools but in the households of the students, though, any student with a household member that was isolating from covid-19 symptoms were excluded from the study. Although, the ‘test-to-release’ study of Young *et al*. (2021) provided positive preliminary data on the effects of LFDs in classrooms, there is further analysis required to understand their use in a variety of infectious environments before promoting ‘test-to-release’ as an effective strategy to prevent transmission.

Mathematical modelling offers a wide variety of techniques to predict the impact of interventions on infection spread without risking anyone’s health [31]. Notably, many different mathematical fields are able to provide predictive results, for example, deterministic and stochastic differential equations [32], machine learning [33], Monte Carlo simulations [34] and queuing theory [35]. For the small number of individuals we are considering a deterministic approach would not be justified [36]. Further, machine learning tools, or continuous time data techniques [37], are not currently viable due to a lack of data. Though infection modelling and analysis has been conducted on the impact of school reopening on local *R* numbers [38]. Hence, a novel, detailed methodology for tracking Covid-19 spread through multiple, linked spatial compartments that is flexible enough to compare a variety of NPIs has been developed and presented here.

We develop an individual-based model of infection spread that splits the susceptible population into local and non-local groups. The algorithm can be used to predict the spread of an infection in many situations, where individuals naturally form groups, or ‘cliques’. We focus on modelling an educational setting; in particular, we seek to predict how testing, isolation and other interventions influence the spread of infection in secondary schools, Further Education (FE) settings (e.g. colleges) and Higher Education (HE) settings, (e.g. halls of residence in a university). Importantly, the local versus non-local distinction can be used to differentiate between short-range airborne transmission due to large droplets and longer-range airborne transmission (aerosol) transmission [39].

Our results are based on current UK data [40]. We also provide the reader access to an online applet in addition to a maintained repository of open source MATLAB code (see Section 2.4) that not only reproduces our results, but can also be easily adapted by the reader to include data which is more accurate and/or specific to their location and needs.

Our results have been presented to the Technical Advisory Group (TAG) of the Welsh Government informing policy on the Covid-19 response in Wales and to the Wales Further and Higher Education Covid-19 Task Group. Our methodology is currently being used to inform policy in relation to the future of students returning to educational environments. Furthermore, our findings have been shared with the Environmental Science TAG Subgroup for use in advising how to open up more general social spaces, such as places of worship. The results have been communicated across the governments of England, Scotland and Northern Ireland, and have informed the wider development of policy planning for the pandemic.

Section 2 presents the assumptions and processes that underlie the algorithm that controls the individual agents (aggregations of pupils, students, etc.) that we are interested in and simulating. Specifically, we clarify how testing and isolation of individuals (agents) is enacted, in sections 2.2 and 2.3. The algorithm is then applied in Section 3 for two settings: (i) a secondary school/FE environment, Section 3.1, and (ii) a hall of residence at a Higher Education establishment, Section 3.2. Critically, within each of these broad applications we consider the effects of how and when LFD testing is applied. Finally, in Section 4 we condense our findings down to simple observations which summarise our findings regarding the applications of LFD testing versus other possible NPIs.

## 2 Computational framework

We have constructed an individual-based stochastic algorithm for infection spread which can be applied to, but not limited to, the Covid-19 pandemic. The code is comprised of independent modules that can be turned on, or off, as appropriate, so that the simulations can explore a variety of realistic situations. For example, we can vary the days and frequency in which testing is conducted. We provide a basic description of the ideas behind the algorithm here with an overview illustrated in Figure 1. A descriptive flowchart of the algorithm is provided in Appendix A.

**Figure 1:**
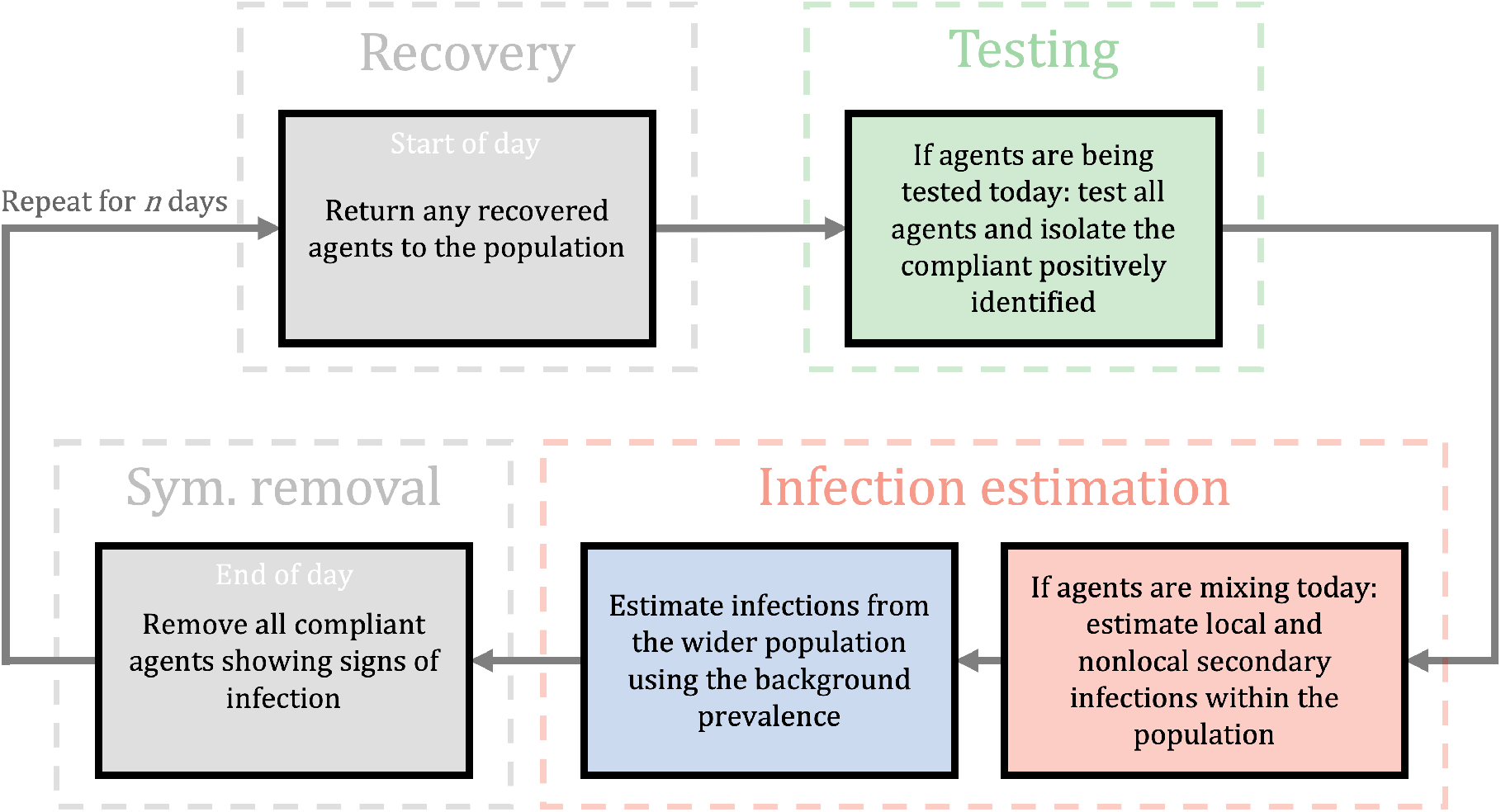
A flowchart illustrating the general iterative process controlling the dynamics of the population over a typical day after the initial simulation setup. Grey boxes represent actions that occur every day whereas the coloured boxes can be independently active/inactive on any prescribed day. A single cycle accounts for a single day. We start each day at the recovery box which returns any isolating agents that have completed their isolation period. If testing is active, the individuals are tested using LFD, agents with a positive test are removed from the population for *t*_*r*_ days. If agents are allowed to mix, then we estimate secondary infections using *R* with context specific scalings (*e*.*g*. NPIs). Next, if external infection sources are being considered, we estimate the number of infections to the susceptible population using the background prevalence. Finally, we remove any compliant symptomatic individuals that display the infection at the end of every day. For a detailed flowchart of the infection algorithm, see Appendix A.

We consider a population of *N* susceptible individuals split into uniform groups of size *N*_*g*_. The parameter *N*_*g*_ is the number of local contacts an individual has and represents the number of individuals that would be instructed to isolated should a member of a group become identified as infectious. If *N*_*g*_ = 1 only an infected individual is isolated upon (i) becoming symptomatic, or (ii) receiving a positive LFD test. Whereas, if *N*_*g*_ = *N* the entire simulated population is isolated upon the identification of an infected individual. Finally, if *N*_*g*_ is any other divisor of *N* then only a subgroup of the population is isolated upon the identification of an infected individual.

As we consider different educational settings we will ascribe different definitions to *N*_*g*_. Namely, in a primary/ junior/ secondary/ FE school setting *N*_*g*_ will represent a ‘table group’. These are individuals who are socially distanced according to current regulations, but are sat on the same table. This is in contrast to the HE case where we consider *N*_*g*_ to be the size of a ‘kitchen group’ (dormitory). These are individuals in shared accommodation in a hall of residence that share the same facilities.

The effectiveness of isolation is modulated by a ‘compliance parameter’, *C*. In the secondary school setting we assume that the school is keeping records of positive tests and has a final say on who is allowed to enter the classroom. Thus, we assume that *C* = 100% and, any student that is symptomatic, or receives a positive test result, is isolated completely for a set amount of time in which they cannot pass on further infections to the simulated population.

However, for the FE and HE cases, information supplied by the Welsh Government’s Head of Policy HE Covid-19 (B. Cradock, 4th Feb 2021, pers. comm.) suggests that students in the HE environment are only 81% likely to comply with an isolation order. Hence, in such cases, after receiving a positive test result, or becoming symptomatic, a HE student is only isolated with a probability specified by the parameter *C*. It is also assumed that a FE student isolates with probability *C* when given cause, due to the open-community approach to education and age range of those studying and FE intuitions.

As discussed above the our decision-making framework accounts for both symptomatic and asymptomatic individuals. Specifically, the percentage of symptomatic individuals is defined as *P*_*s*_. At the point of infection each individual is either given a symptomatic, or an asymptomatic flag based on this percentage. Asymptomatic individuals are able to infect other susceptible individuals but will not isolate unless tested, whereas symptomatic individuals are assumed to follow public health guidance and follow self-isolation guidance at a rate determined by the compliance parameter. Symptomatic individuals who choose not to be tested are treated as asymptomatic. A non-exhaustive list of real-world infection events and model interpretation is supplied in Table 3 in Appendix A.

The timescale of the algorithm is one day. For every day we specify whether individuals are to be tested and whether they are able to mix. These scenarios are independent and, thus, we control whether neither, one, or both scenarios occur. For example, because we are modelling a school, or college location, we currently specify that secondary and FE students do not mix on a weekend and, equally, no testing happens on a weekend. Whereas in the halls of residence related to the HE simulations mixing and testing can occur every day.

As seen at the beginning of the flowchat in Figure 1, during any day that testing is applied all individuals that are not isolating are tested. The testing phase occurs before the mixing phase. Thus, any individual triggering a positive result is isolated before they can infect others. Critically, we currently do not include infection transmission once individuals are isolating as we suspend any isolating agent from the simulation for *t*_*d*_ days. Specifically, in a secondary school, if the whole class is isolating no further infections appear. This assumption is reasonable in the secondary and FE cases as the students are physically isolated from one another. However, in the HE case, further infections may occur within isolated flats. Thus, when we compare the isolating individuals across those that are infected and those that are not we only quote this ratio to be correct at the point of isolation, because, as mentioned, further infections within an isolating subgroup are not tracked but are likely to occur, see Section 4.2.

Due to the algorithm being able to contend with subgroups within a population we have effectively added two compartment spatial complexity to the infection model. Namely those within a group are considered ‘local’ contacts and those outside of a group are considered ‘nonlocal’ contacts. Because of this addition of spatial complexity, the basic reproduction number, *R* is split into two *R* numbers:

1. a local number, *R*_*l*_, that measures the expected number of secondary infections within a subgroup that has an infected person; and
2. a non-local number, *R*_*n*_, that measures the expected number of secondary infections that occur due to infectious people in other subgroups.

Critically, the basic reproduction number, *R*, can be estimated from data and *R*_*l*_ and *R*_*n*_ are defined such that *R* = *R*_*l*_ + *R*_*n*_. Specifically, we define a ratio of local to non-local infections, *c*_*l*_ : *c*_*n*_, and, thus, *R*_*l*_ = *c*_*l*_*R/*(*c*_*l*_ + *c*_*n*_) and *R*_*n*_ = *c*_*n*_*R/*(*c*_*l*_ + *c*_*n*_).

Defining the local to nonlocal infection ratio, *c*_*l*_ : *c*_*n*_, depends on how we assume the virus spreads. For example, in a secondary school environment, if there is a lot of movement, talking, and intergroup mixing then we might expect there to be no difference between the number of infections within groups to infections between groups, in which case *c*_*l*_ = *c*_*n*_. However, if a good social distancing policy is implemented, we would expect the number of nonlocal infections to be lower than the number of local infections, *i*.*e. c*_*n*_ < *c*_*l*_.

Explicitly, for the secondary school simulations, we are choosing to inform *R*_*l*_ and *R*_*n*_ through the steady state level of spread of airborne particles [41]. In [41], the concentration of airborne particles in a classroom is determined by solving an advection–diffusion–reaction equation (see Section 2.1). Applying the same methodology here, we evaluate the local contagion concentration, *c*_*l*_, at 2 metres directly downwind from an infected individual in the centre of the room. This represents the worst-case scenario for the amount of contagion that a susceptible person would receive if there was an infected individual in their subgroup and social distancing was strictly observed. Further, we evaluate the nonlocal contagion concentration, *c*_*n*_ at 4 metres distance in the direction orthogonal to the airflow from an infected individual. This represents the mean average scenario for the amount of contagion that a susceptible person would receive if there was an infected individual in another subgroup. Note that here the ratio *c*_*l*_ : *c*_*n*_ is fixed, with the values taken from the airborne simulations at 5 hours, which is consistent with the day time scale, on which we are working. However, this ratio should actually be evolving as predicted by [41].

In the HE case, where we consider the infection spreading through a Halls of Residence we choose to define this ratio based on the compliance of the individuals to social distancing policy. Specifically, we consider two cases. The first is the ‘poor social distancing’ where residents from all flats mix, *i*.*e*., the local and nonlocal infection numbers are equal (*c*_*l*_ : *c*_*n*_ is 1:1, or *R*_*l*_ = *R*_*n*_). The second case considers ‘enhanced social distancing’, where on average, 5 out of 6 flat residents are only socialising with members of their same flat, whilst 1 in every 6 people still mix with other flats even when strict social distancing measures have been advised. Thus, in the enhanced social distancing case, the ratio of local to nonlocal infections is 5 : 1. This value was estimated to agree approximately with the isolation compliance probability of *C* = 81%. However, we did not simply choose a ratio of 4 : 1, which would match this compliance, because the ratio of 5 : 1 is more interpretable when the flat occupancies are of size 6, or 12, which are prototypical HE dormitory capacities in the UK. Namely, on average, there is one, or two, poorly compliant individuals in the flats, respectively.

Alternatively, instead of splitting *R* into local and nonlocal contributions there is the future possibility of using research focused on simulating airborne particle spread to specify *R*_*l*_ and *R*_*n*_ directly [41], thus providing a mechanistic way of understanding *R*. This could be encompassed into our simulation, although we would need to reduce the resolution of time within the model from hours to day and therefore inducing greater computational costs.

We now consider the blue and red sections of Figure 1, which are the infection stages. The blue section controls external infections, namely infections that occur outside of a school, or Hall of Residence, as appropriate. In the secondary school scenario, we assume that these happen at the end of the day due to socialising after school, which is why the red section feeds into the blue section. These infections occur at the local prevalence rate, *I* [42] can be turned on, or off, depending how well we believe our system is isolated from the outside world. Equally, isolating the system allows us to consider how the system influences itself under any intervention.

The red sections of Figure 1 control how secondary infections are generated due to mixing between individuals. During any day where the students can mix then any individual that is:

- infected;
- infectious; and
- not isolated

can infect other people. Once a person becomes infected three delay clocks are attached to each of them. The three delays [43], *t*_*i*_, *t*_*d*_ and *t*_*r*_, represent:

- the time between becoming infected to becoming infectious, *t*_*i*_ = 3 days;
- the time between becoming infected to becoming detectable by a LFD test, *t*_*d*_ = 5 days; and
- the time between becoming infected to recovery, *t*_*r*_ = 10 days.

Any recovered person is assumed to be non-infectious and immune to further infections. Currently, these delays are fixed, but these could easily be made variable. Note that the recovery and isolation timescales are independent. Thus, once an individual is discovered to be infected they are isolated for 10 further days, no matter where they are within the infection timeline. For example, if an individual is found on the 9th day of their infection they have to isolate for another 10 days, even though they would recover the next day.

Finally, we assume that the tests being used are Lateral Flow Devices, LFD, which are assigned to have a false positive probability, *P*_*fp*_ and a false negative probability of *P*_*fn*_. Throughout the simulations, we will vary *P*_*fn*_ but keep the false positive probability fixed to *P*_*fp*_ = 0.003, in agreement with existing LFD studies [19, 22, 44].

### 2.1 Interventions

In order to quantify the effects of environment activity, masks and ventilation on transmission, we employ a previously developed advection-diffusion-reaction equation to estimate the concentration of infectious particles within a bounded 3D space [41]. The contagion density model extends the Wells-Riley model of airborne infectious diseases and is of the form

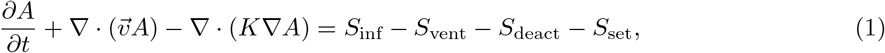

where *A* is the concentration of airborne infectious particles (particles/m^3^), *t* is the time (s). In addition, *S*_inf_ represents the source of infectious particles (i.e. the infectious people); *S*_vent_, *S*_deact_, *S*_set_ are sink terms representing the removal of particles by three factors: the ventilation system, biological deactivation, and gravitational settling. Specifically, equation 1 models viral particles advected by airflow, diffused due to turbulence, emitted by infected people and removed due to the room ventilation. The equation also includes terms that model the biological inactivation of the virus and gravitational settling. One asymptomatic or presymptomatic infectious person who breathes or talks was considered, who may wear a mask. Equation 1 has a semi-analytic solutions that allows for the swift quantification of the effect of NPIs on infection transmission.

Additionally, we estimate the infection risk (probability of infection), *I*_*A*_, using the dose-response formula

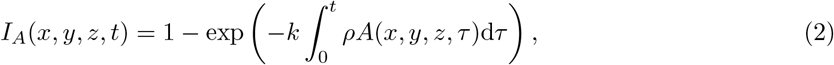

at any location within the 3D space, where *k* is an constant that depends on the infectiousness of the virus, and *ρ* is the average breathing rate. Hence, the integral calculates the number of infectious particles inhaled by the susceptible person. Further details on airborne infection estimation can be found in [41]. The epidemiological *R* value can be modified by additional interventions mitigating this transmission, for example, masks [45], ventilation [46] and portable air purifiers [47]. Based on realistic scenarios and recommended ventilation settings by ASHRAE (the American Society of Heating, Refrigeration and Air-Conditioning Engineers) [48], we considered four ventilation settings:’very poor ventilation’ (ACH= 0.12), ‘poor ventilation’ (ACH= 0.72), ASHRAE pre-pandemic ventilation (ACH= 3.00) and ASHRAE pandemic-updated ventilaton (ACH= 6.00). Using equations 1 and 2, we estimate the ratio of particle concentrations and infection probabilities between local and nonlocal individuals using a prototypical classroom environment for each of these ventilation scenarios (see Appendix C). Specifically, we assume that masks halve the emission rate of particles [49] and consequently reduces the *R* value by the same amount [41]. In addition, we also assume a good ventilation setting with ACH equal to 6.0 which yields a 3.5-fold decrease in *R*. The combined influence of masks and ventilation is assumed to be multiplicative, in which case *R* is reduced 7-fold [41].

Improving ventilation also decreases the local to nonlocal infection ratio, as better ventilation promotes better mixing of the air in the room with a reduced contagion density [50]. The usage of masks has the opposite localisation effect by preventing contagion from propagating significant distances [51]. The combination of these effects in classroom environments can be found in Appendix C. Throughout all simulations conducted here in this study, we assume that the individuals will have low activity (intermittent talking) within the classroom environments and apply the best and worst-case ventilation scenarios. Therefore we have the resultant local to nonlocal infection ratios by from 1 and 2 in Table 1.

**Table 1:** A summary of infection parameters generated by equations 1 and 2 for individuals under low activity in a classroom environment. For details on parameter estimations, see Appendix C.

| Environment scenario | Effective $R$ | Local to nonlocal infection ratio ($c_l : c_n$) |
| --- | --- | --- |
| Very poor ventilation + no masks | $R$ | 3.98:1 |
| Very poor ventilation + masks | $R/2$ | 5.20:1 |
| Good ventilation + no masks | $2R/7$ | 1.47:1 |
| Good ventilation + masks | $R/7$ | 1.51:1 |

In the Halls of Residence scenarios ventilation capabilities cannot be easily changed without significant cost - solutions like portable air purifiers could be considered [52]. Further, masks are usually not worn whilst individuals are in their own flats. Thus, we assume that the enacted interventions are reducing the number of non-local contacts an individual has, following guidance from the university and the government. Specifically, for the HE scenarios we do not change *R* directly as interventions are introduced, we change the ratio of the number of local to non-local infections. In the so-called ‘poor social distancing’ scenario residents from all flats mix and, thus, the local and non-local infection numbers are equal, that is *c*_*l*_ : *c*_*n*_ is 1:1. In the ‘enhanced social distancing’ scenario individuals are encouraged to isolate as much as possible, and they, more likely, socialise only within their flat group (‘kitchen group’). This reduces the number of non-local infections, compared to the local infections; we thus, take the ratio *c*_*l*_ : *c*_*n*_ to be 5:1/5, for reasons stated above, in Section 2.

At the time of conducting this study, vaccination of people aged 12-16 had not been implemented in the UK [53]. Therefore, we do not consider vaccinations as a method of transmission mitigation in both the secondary schools and HE simulations, particularly to characterise the infection landscape of the worst-case scenario and to isolate the use of NPIs in educational environments. We acknowledge that vaccination policy may change in the future and so immunity via vaccination or recent infection has been implemented into the online simulator of the model (https://bit.ly/CV19_INTER_IBM) to allow for up to date simulation results (see Appendix B). Moreover, by including the immunisation into the applet, the model may be applied to regions where youth vaccination is occurring.

### 2.2 Testing regimes

Alongside varying the false negative probability of the LFD we consider a variety of different testing and isolation scenarios. The base case against which we compare all other cases is the no testing and no isolation of contacts case. Namely, the disease is simply allowed to spread through a class, or Halls of Residence without impedance.

Noting that in the cases modelled, only non-isolating individuals use LFD devices we can alter the testing regime for the population from weekly to daily. Further, we can alter the event that causes testing to occur. Either testing occurs on a fixed regime, or alternatively, no testing is applied until a first symptomatic person is found, that is, there exists an agent showing symptoms that day. Once a symptomatic individual is found, testing is applied through a weekly, or daily regime.

Other “reactive” testing regimes can be included (*e*.*g*., you always test after a symptomatic is found, rather than just moving to a fixed regime), but the “no testing” to “daily testing” range provides the extreme limits in which all other possible testing scenarios must fall. In Section 3 we will see that these extreme testing scenarios provide limits over a small range of results and, thus, all other proposed testing regimes must fit within these limits. Hence, understanding only the limiting cases provides us with enough knowledge to understand all cases.

As a specific example, in the secondary school case, we are particularly interested in the case of “test-to-release”. Namely, it was previously the case that if an infected individual was found then their close contacts would be isolated too, in this case this would be their table group (see Figure 2(c)). However, current guidance [5, 6, 7] is that a negative LFD result could release individuals in the table group that test negatively. Under our simulation this would conform to the individual isolation case. Namely, only the individual is forced to isolate, whilst everyone else is tested before they are then allowed to mix (see Figure 2(b)).

**Figure 2:**
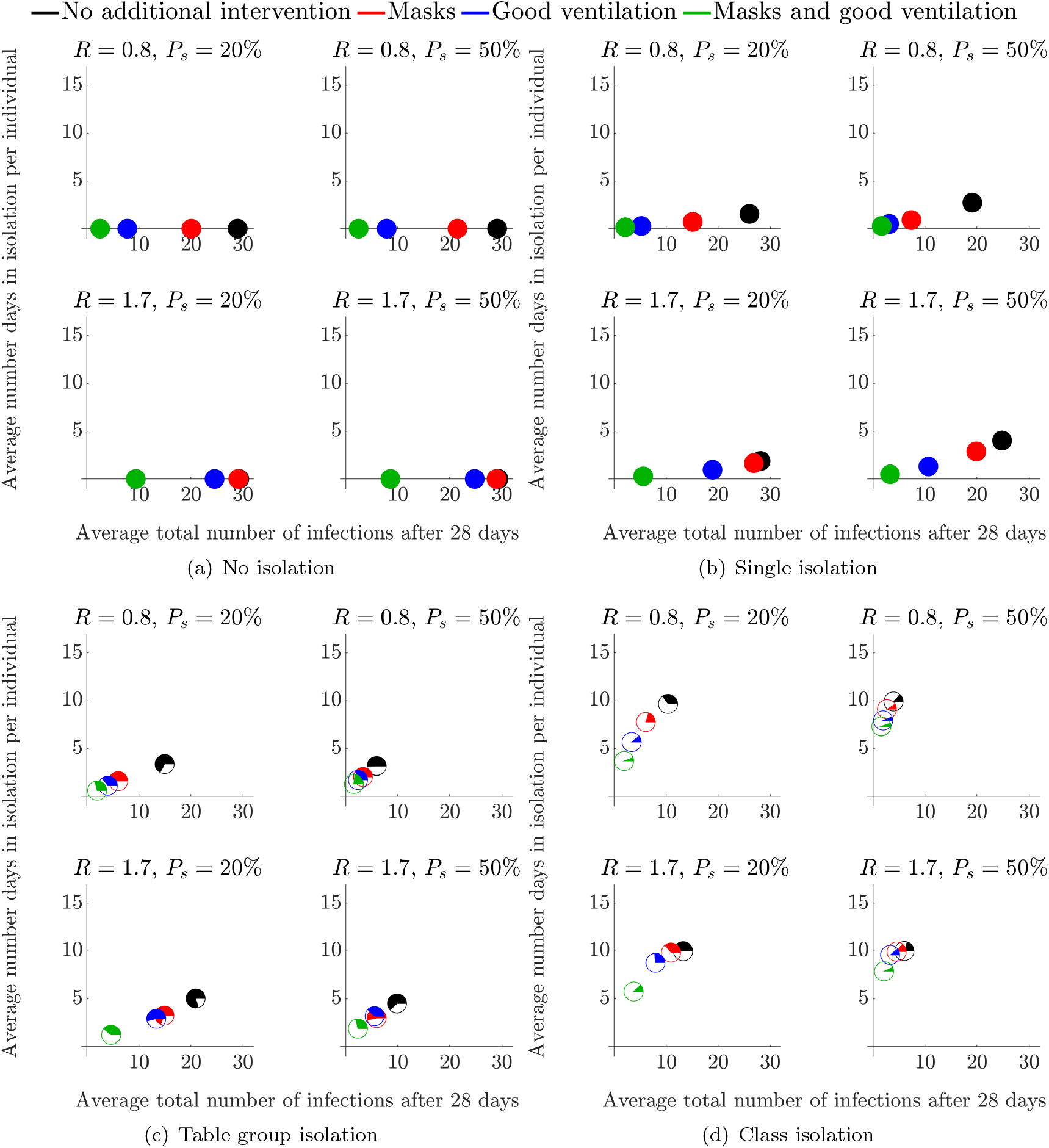
Mean number of infected students and average number of days absent per individual when no testing is employed, over a variety of isolation strategies and interventions in a secondary school environment. Each symbol represents a different intervention, see legend at the top of the figure for details and see Section 2.1 for a description of how each intervention influences the parameter values.

In the HE setting we compare the efficacy of the various testing strategies at two time points during a term. In the first scenario, called the ‘start of term’ scenario, the first simulated week occurs prior to students returning to the Halls of Residence. In this initial week students do not mix, but any infected student will incubate their infection. The next three weeks are simulated under the assumption that the students are back in their halls and able to mix daily. The second scenario, called the ‘middle of term’ scenario, considers a period post student arrival, where the individuals are mixing each day (including weekends) for four weeks.

### 2.3 Isolation regimes

As mentioned in Section 2.2 we initially consider the case in which no isolation occurs. This situation can then be compared against:

- isolating individuals due to the individual being symptomatic, or receiving a positive test result;
- isolating subgroups due to at least one individual in the subgroup being symptomatic, or receiving a positive test result;
- isolating an entire population due to at least one individual being symptomatic, or receiving a positive test result;

Note that the positive test results leading to an isolation can be either true or false positives.

Throughout the simulation we keep track of the number of people who are isolating. Further, the scatter points illustrated in Section 3 are all pie charts that represent the ratio of infected people who are isolating (white part of the pie chart) versus the number of people who are isolating, but healthy (colour part of the pie chart).

### 2.4 Scenario parameters

To elucidate the robustness of the LFD testing strategies in various infectious environments, we simulate the mathematical model across a variety of parameter values. A best-case scenario would be when there is: a low background prevalence; a low *R* number, and a low probability of false negatives, *P*_*fn*_. In contrast, the situation becomes worse when any of these parameters are increased. For each parameter we choose two extreme values (see Table 2) and simulate over all combinations of these values. In addition to these global parameters, we provide a summary of context specific parameter values and parameter definitions in tables 4 and 5 in Appendix A. Note that the *R* number, as referenced in tables 2 and 5, is the local background *R* number, that is parameterised using the National *R* number estimates for our best- and worst-case scenarios [54]. The actual numbers that apply in School or University environments are anticipated to lie between these extremes and may depend on whether transmission between children, adolescents and young adults differ substantially from aggregate National *R* numbers [55]. Clearly local strategies such as increasing ventilation or mandating mask use may already contribute to the lower estimate of the National *R* number. Nonetheless, we anticipate that the modelled NPIs will apply between the two *R* numbers suggested.

**Table 2:** Table of parameter values used in all education environments simulations.

| Parameter | Definition | Best-case parameter values | Worst-case parameter values |
| --- | --- | --- | --- |
| Probability of false negatives, $P_{fn}$ | Likelihood that a Covid-19 test does not identify an infectious individual. | 0.2 | 0.5 |
| R number, $R$ | The average number of secondary cases we would expect an infected student to generate prior to locality affects. | 0.8 | 1.7 |
| Background prevalence, $I$ (%) | Additional probability that more than one person is infected. | 0.5 | 2.0 |

**Table 3:** A table representing real-world infection events as interpreted by the model framework presented in this study.

| Real-world event | Infection classification | Infectious to the population at post event | Maximum number of days infectious prior to event (days) | Maximum number of days infectious post event (days) | Isolation status post event | Susceptible status post event |
| --- | --- | --- | --- | --- | --- | --- |
| Presymptomatic student attending school | Asymptomatic | ✓ | 0 | $t_d - t_i$ | ✗ | ✗ |
| Non-compliant symptomatic student attending school | Asymptomatic | ✓ | $t_d - t_i$ | $t_r - t_d$ | ✗ | ✗ |
| Symptomatic student with a true-positive test result compliant with isolation policy | Symptomatic | ✗ | $t_r - t_i$ | 0 | ✓ | ✗ |
| Symptomatic student with a false-negative test result compliant with isolation policy | Asymptomatic | ✓ | $t_r - t_i$ | 0 | ✓ | ✗ |
| Asymptomatic student with a true-positive test result compliant with isolation policy | Asymptomatic | ✗ | $t_r - t_i$ | 0 | ✓ | ✗ |
| Asymptomatic student with a false-negative test result | Asymptomatic | ✓ | $t_r - t_i$ | $t_r - t_i$ | ✗ | ✗ |
| Non-infected student with a false-positive test result from random testing | Non-infected | ✗ | 0 | 0 | ✓ | ✓ |
| Non-infected student with a true-negative test result from random testing | Non-infected | ✗ | 0 | 0 | ✗ | ✓ |
| Asymptomatic student in close contact with a positively identified infected student | Asymptomatic | ✗ | $t_r - t_i$ | 0 | ✓ | ✗ |
| Non-infected student in close contact with a positively identified infected student | Non-infected | ✗ | 0 | 0 | ✓ | ✓ |

**Table 4:** A summary of parameter values used for secondary school and HE simulations. See Section 2.2 for information on various testing regimes.

|  | Number of agents | Testing | Agent mixing | Wider pop. infections | Isolation group sizes | Probability of symptomatic | Probability of compliance |
| --- | --- | --- | --- | --- | --- | --- | --- |
| Secondary school (no testing) | 30 | ✗ | Weekdays | ✗ | 1, 5 & 30 | 0.2 & 0.5 | 1.0 |
| Secondary school (with testing) | 30 | ✓ | Weekdays | ✗ | 1, 5 & 30 | 0.2 & 0.5 | 1.0 |
| Halls of residence (start of term) | 204 | ✓ | 1 week isolated followed by 3 weeks continuous mixing | ✗ | 6 & 12 | 0.4 | 0.8 |
| Halls of residence (middle of term) | 204 | ✓ | Everyday | ✗ | 6 & 12 | 0.4 | 0.8 |

**Table 5:** A table of model parameters with their associated definitions.

| Parameter | Definition |
| --- | --- |
| Probability of false-positive, $P_{fp}$ | The likelihood that a LFD test identifies a non-infected agent with a positive result. |
| Probability of false-negative, $P_{fn}$ | The likelihood that a LFD test does not identify an infected agent with a positive result. |
| R number, $R$ | The average number of secondary cases expected from an infected agent to generate prior to locality affects. |
| Local R number, $R_l$ | The proportion of R that accounts for the number of secondary infections within a subgroup of the population. |
| Nonlocal R number, $R_n$ | The proportion of R that accounts for the number of secondary infections to those outside the infectives subgroup. |
| Background prevalence, $I(\%)$ | The rate at which the wider population introduces an infection to an agent. |
| Probability of compliance, $C$ | The likelihood that an agent is compliant with testing and isolation policies. |
| Probability of symptomatic, $P_s$ | The likelihood that an infected agent will become symptomatic. |
| Total population size, $N$ | The total number of agents in the simulation. |
| Subgroup size, $N_g$ | The number of agents that make up a subgroup within the total population. |
| Infectious time, $t_f$ | The time between becoming infected to becoming infectious. |
| Detectable time, $t_d$ | The time between becoming infected to becoming detectable by a LFD. |
| Recovery time, $t_r$ | The time between becoming infected to recovery from infection. |

### 2.5 Code availability

Our intention is to illustrate our code in a number of educational settings, but our algorithm can be used to predict infection numbers across variety of other locations. Equally, we provide one interpretation of how interventions influence the outcome. The actions and interpretations of the interventions are completely arbitrary and, thus, the algorithm can be exploited to provide predictive results including a new variety of interventions, so long as the user has a clear idea how the interventions influence the input parameters.

Moreover, our interest is to investigate a wide space of reasonable parameter values to illustrate how diverse our results can be. As parameter estimates become better over time, due to more data being collected, we may want to return to our simulations and run the computation under specific parameter values that represent our current knowledge of a system. Alternatively, in line with the above comments about extending the interpretation of our code, the parameters will also have to be reinterpreted within the new scenarios.

To account for this diversity of use we provide open access to an online simulator of the model presented here which is dedicated to secondary school and FE environments (https://bit.ly/CV19_INTER_IBM). The online applet allows the user to both reproduce the results of this study and to design new infection scenarios with current infection data without the requirement of coding ability or software. Appendix B contains information on the online COVID-19 Intervention Simulator, usage and features. In addition, all source code for the model and online applet is stored and maintained at https://github.com/joshwillmoore1/COVID-19_Intervention_IBM. The results shown in this study were run and tested on MATLAB 2020b and the online applet was developed using R v4.0.3. We hope that by developing a user-friendly simulator of our model and making the source code openly available allows others to expedite their response to the pandemic through predicting which of their intervention options is the best.

## 3 Results

The algorithm is able to output an immense amount of data. Specifically, we track the infection, isolation and symptomatic status of every individual with a day-time resolution. Further, due to the stochastic nature of the algorithm, the code is run 1000 times for each scenario, which provides us with an understanding of the sensitivity of our estimates. Thus, although many daily statistics are available as outputs from the code, we visualise only the average total number of infections versus the average number of days isolating per individual.

Note that the speed of the simulations is mainly determined by the size of *N*, since we fix the number of repetitions. Thus, for the FE classes, which are of size *N* = 10, 1000 simulations takes approximately 9.9 seconds to complete. However, in the case of the Halls of Residence, where *N* = 204 the results take approximately 40.3 seconds^1^. Sweeping over the 5-dimensional parameter space required to analyse the best and worst-case scenarios, simulation times scale linearly, achieving a maximum simulation time of approximately 4 hours to complete in the HE settings.

As mentioned in Section 2.3 we present a number of Figures that use pie charts to track numbers of infected or healthy individuals isolating at points that are scattered in the space of average ‘infections’ versus ‘days of isolation per individual’. Each pie chart represents a different intervention. The intervention is encoded in the colour and transparency of the pie chart. The coloured part of each pie chart represents the proportion of ‘correct’ isolations, *i*.*e*. isolations of infected people, whereas the white section represents the proportion of incorrect isolations, *i*.*e*. isolations of healthy people. Thus, when seeking to optimise NPIs, we may independently assess overall infection rates and approaches that minimise the isolation of uninfected individuals.

Overall, we assume policy makers are primarily interested in minimizing the total number of infections and the number of days spent in isolation. Thus, we look to strategies that produce points near to the origin, (0,0) with the lowest proportion of uninfected individuals isolating (coloured section of pie charts). When these two goals are in conflict, it would be assumed that minimising the total number of infections would have priority.

### 3.1 Secondary school environments

In this case we set the class size to be *N* = 30 and consider *N*_*g*_ = 1, 5 and 30. Thus, apart from the case where no isolation occurs, we will either be isolating: only an infected individual, an infected and their table group, or the entire class, respectively (see Table 3 for our interpretation of real-world events in the modelling framework).

As a base case for investigating the efficacy of testing we, first, run the model without testing included. Thus, the parameter *P*_*fn*_ is irrelevant. Thus we focus on varying the level of symptomatic prevalence, *P*_*s*_ between 20% and 50%. Having a large population of asymptomatics, potentially causes problems when we run a reactive testing strategy, rather than a fixed testing strategy (see Section 3.1.2) [56, 57]. When testing is considered, we fix *P*_*s*_ = 20%, which provides the worst-case scenario of 80% of the infected population not presenting symptoms.

Equally, from simulating all combinations of the best- and worst-case parameters as discussed in Table 2 we note that reducing *R* reduces the average total number of infected people and the average number of days isolation better than reducing the false negative probability, *P*_*fn*_. Noting that this observation remains true over all simulations we suppress the data from mixed simulation for clarity of discussion and only present the best- and worst-case parameter values.

The simulations are run over 28 days and we assume that the simulation starts on a Sunday, see Figure 6 for an explicit description of simulation initialisation. Defining the starting day is important as we assume that testing and mixing can only occur during weekdays, not weekends.

In the context of a secondary school, the Head Teacher has the authority to remove any student displaying symptoms from the classroom. In addition, as the school will oversee testing and recording of results, we set the percentage of student compliance to 100%. From data not shown, we note that varying compliance and background infection rates, within realistic limits, (*i*.*e*., 60 ≤*C* ≤ 100 and 0.5 ≤ *I* ≤ 2) does not significantly influence the forthcoming insights, thus, we have suppressed this output. Reducing the local prevalence rate *I* to 0.5% from 2% has an insignificant effect on the number of infections within the classroom due to the population size being small (*N* = 30). Namely, we would have required at least a prevalence rate of *I* = 3.3% to infect at least one student per simulation by wider population transmission. Although changing the compliance has the obvious influence on the quantitative results the qualitative results remain the same from the perspective of intervention efficacy. Hence, in the following figures all simulations have a background infection prevalence of *I* = 2% and a compliance of *C* = 100%.

#### 3.1.1 Without testing

Figure 2 presents the simulations under the assumption of no LFD testing. Thus, in the cases where isolations occur, isolations are only able to occur when a student becomes symptomatic (3 days post infection) and/or when those who have been exposed to possible infection are asked to isolate.

Figure 2(a) presents the worst-case scenario where individuals are exposed to a presymptomatic infectious individual under circumstances when school contacts are not isolated. Thus, if there are no interventions then eventually everyone becomes infected within a four-week period (i.e. the black pie chart is always at 30 on the x-axis). Note that, since there are no isolations, we have not isolated any healthy people, thus, the pie charts are fully coloured. Equally, due to having no isolations, even from students showing symptoms, there are no absences, thus, all pie charts lie on the x-axis. In Figure 2(b) only the single symptomatic infected individual isolates, whereas in figures 2(c) and 2(d) the school has been notified and either the table group, or entire class, respectively, has been asked to isolate, as well.

We note from Figure 2(a), which is true over all subfigures and subplots that the level of intervention is increased fewer people become infected. Moreover, although having both masks and ventilation is the best policy, increased ventilation is the better single intervention, because of the substantial impact of diffusing high contagion regions and therefore mitigating risk of infection by removing the particles from the air. This is seen through noting that the green and blue pie charts are much more left in all cases than the red and black pie charts.

Comparing rows and columns in each subfigure shows that increasing *R* increases the average number of infected individuals as expected. An increase in the percentage of individuals who display symptoms (*P*_*s*_) reduces the total number of infected people symptomatic individuals self-isolate (compare figures 2(a) and 2(b)). As an indirect consequence, a reduced number of infections feeds through into a lower average number of days in isolation per individual (Figure 2(b)). A reduction in *R* similarly leads to a reduction in both infection and isolation metrics.Critically, although influencing both parameters is beneficial for our purposes, we are unable to physically influence *P*_*s*_ as this is an intrinsic property of the infection. Noting that these observations of altering *R* and *P*_*s*_ remains true over all simulations then in future figures we only focus on simulations where *R* is varied as this is the parameter that NPIs can influence.

Another result that is consistent across all our simulations and across the secondary, FE and HE scenarios is that isolating larger subgroups of the population is one of the surest ways of reducing the average total number of infections. The contribution of contact isolation becomes greater when good ventilation and mask use is not in place. However, this strategy increases both the number of days in isolation and the number of healthy individuals isolating. Specifically, as the isolation group size is increased from the individual, to the table group and, finally, to the entire class (figures 2(b)-2(d), respectively) the pie charts move left, up and the white sector becomes larger.

#### 3.1.2 With testing

Figure 3 represents the same intervention strategies as those shown in Figure 2, however, we consider multiple testing scenarios. As mentioned, testing frequency is denoted by pie chart transparency. Namely, as the testing frequency is increased the pie charts become more transparent. Note that the no testing data from Figure 2 has also been included as the opaque pie charts, to allow for comparison against the base case.

**Figure 3:**
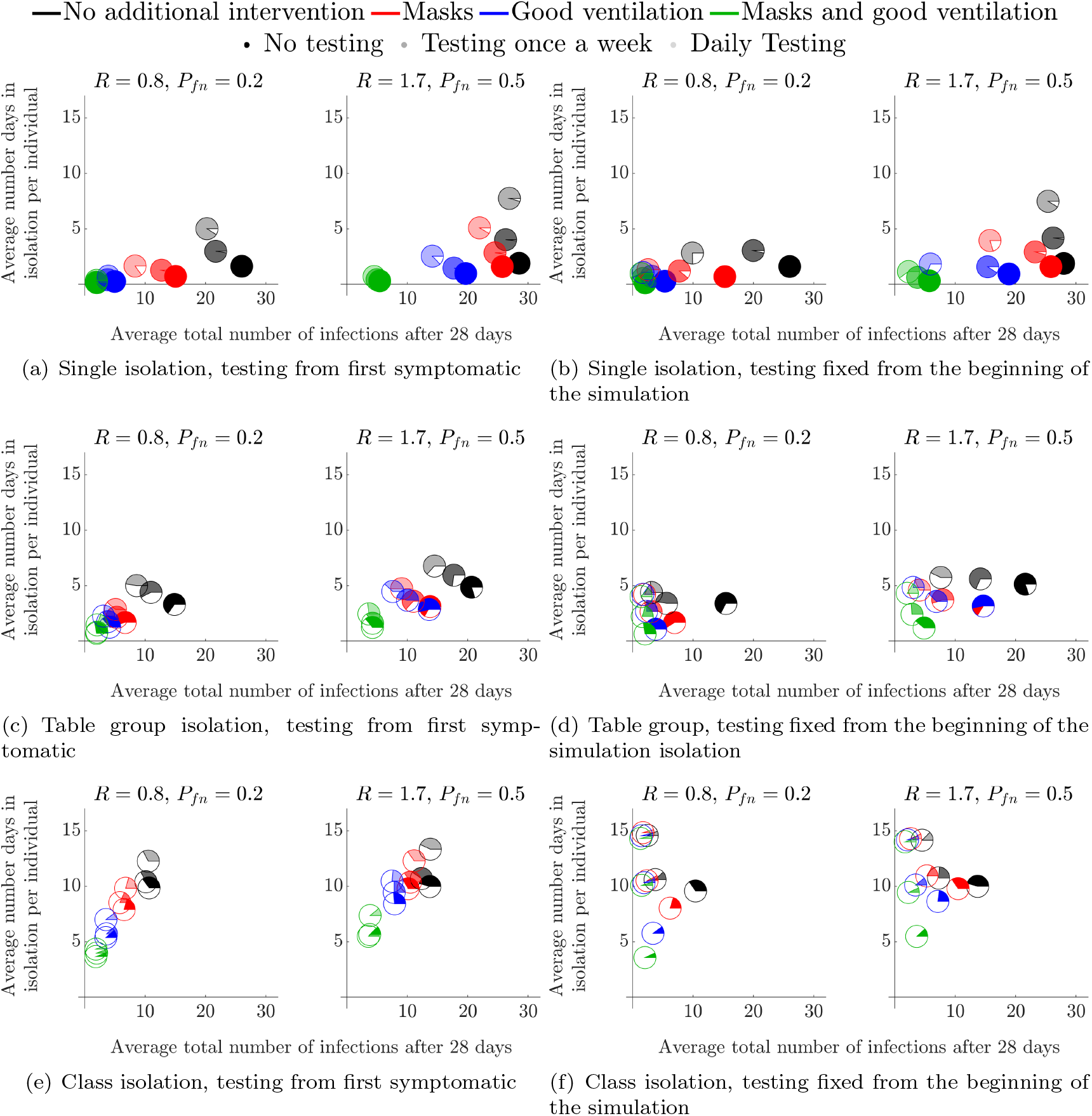
Mean number of infected students and average number of days absent per individual when testing is employed, over a variety of isolation strategies and interventions in a secondary school environment. Each symbol represents a different intervention, see legend at the top of the figure for details and see Section 2.1 for a description of how each intervention influences the parameter values.

The simulations appearing in the left column of Figure 3 all assume that testing only occurs after a first symptomatic individual appears (“reactive” testing), whereas the right column has a fixed testing strategy from the beginning of the simulation. Clearly, the outcome difference between these two strategies is minimal, although they may not differ for cost of implementation. The reason is that, even at a symptomatic (to asymptomatic) prevalence of 20%, a symptomatic individual is generated extremely quickly through secondary infections, which cause the testing strategies to begin almost as soon as the fixed strategies (random testing for asymptomatic students).

The overall trends mentioned in Figure 2 remain the same when comparing the different levels of testing, namely: reducing *R* is the best way of reducing further infections; ventilation is better than masks; and isolating more people greatly reduces the overall number of infections but increases the number of absences and the number of healthy people being isolated.

Notably, we see that the addition of weekly, or daily tests does reduce the number of overall infections, but more tests lead to more absences as more cases are found earlier in the infection cycle (the pie charts move left and up as they become more transparent). Critically, although testing does help, we see that reducing *R* is much more important, particularly due to the time-lag between being infectious and being detectable by LFD (*t*_*d*_ > *t*_*i*_) and so testing will never be able to fully remove infections from the population under our assumptions. As a result, simply wearing a mask and having no testing is as good as daily testing without a mask in a majority of the test scenarios (the opaque red pie chart is always lower and left of the most transparent black pie chart).

When considering ‘test-to-release’ strategies, we note that infection rates were lower when pupils are asked to isolate if they have been in contact with infected individuals. Namely, in Figure 3(e) where *R* = 1.7 and *P*_*fn*_ = 0.5, the black, red, blue and green rates (solid colour; no testing) at 14, 11, 7 and 3 respectively. By contrast the equivalent numbers for pupils who are tested and released back to the School Population in a ‘test-to-release’ strategy (in Figure 3(a) *R* = 1.7 and *P*_*fn*_ = 0.5, the black, red, blue and green rates (transparent colour; daily testing) are 28, 26, 20 and 5. This would suggest that, if the primary policy goal was to minimise infections, that ‘test-to-release’ would increase infections by comparison with either the table-isolation or whole-class isolation strategies.

### 3.2 Higher Education environments

The cohort size in this case is the population of a typical Halls of Residence, *N* = 204. These individuals are separated into flats, or ‘kitchen groups’ of size *N*_*g*_ = 6, or *N*_*g*_ = 12. We fix the symptomatic prevalence to be *P*_*s*_ = 40% and the compliance to be *C* = 81%.

Here, we mainly focus on trying to understand the influence of the flat size on the results and timing of the test. Specifically, see Section 2.2, where we discuss the start of term and middle of term testing scenarios.

#### 3.2.1 Start of term

The start of term simulations starts a week prior to student mixing. Hence, all initially infected individuals are potentially detectable by the first administered LFD test in both strategies. Therefore, the lag time between being exposed to the virus, being infectious and being detectable has no effect on the initial transmission within the student population. In this scenario we consider the effect of a test prior to returning to the Halls of Residence. Namely, in Figure 4, the blue pie charts represent the case in which students are tested two days prior to arrival (Friday of week 1), followed by weekly tests, every Monday, starting on the day of arrival (Monday of week 2). The red pie charts represent the case where there is no test before returning. However, weekly tests are administered, every Monday, starting on the day of arrival (Monday of week 2), as in the blue pie chart case.

**Figure 4:**
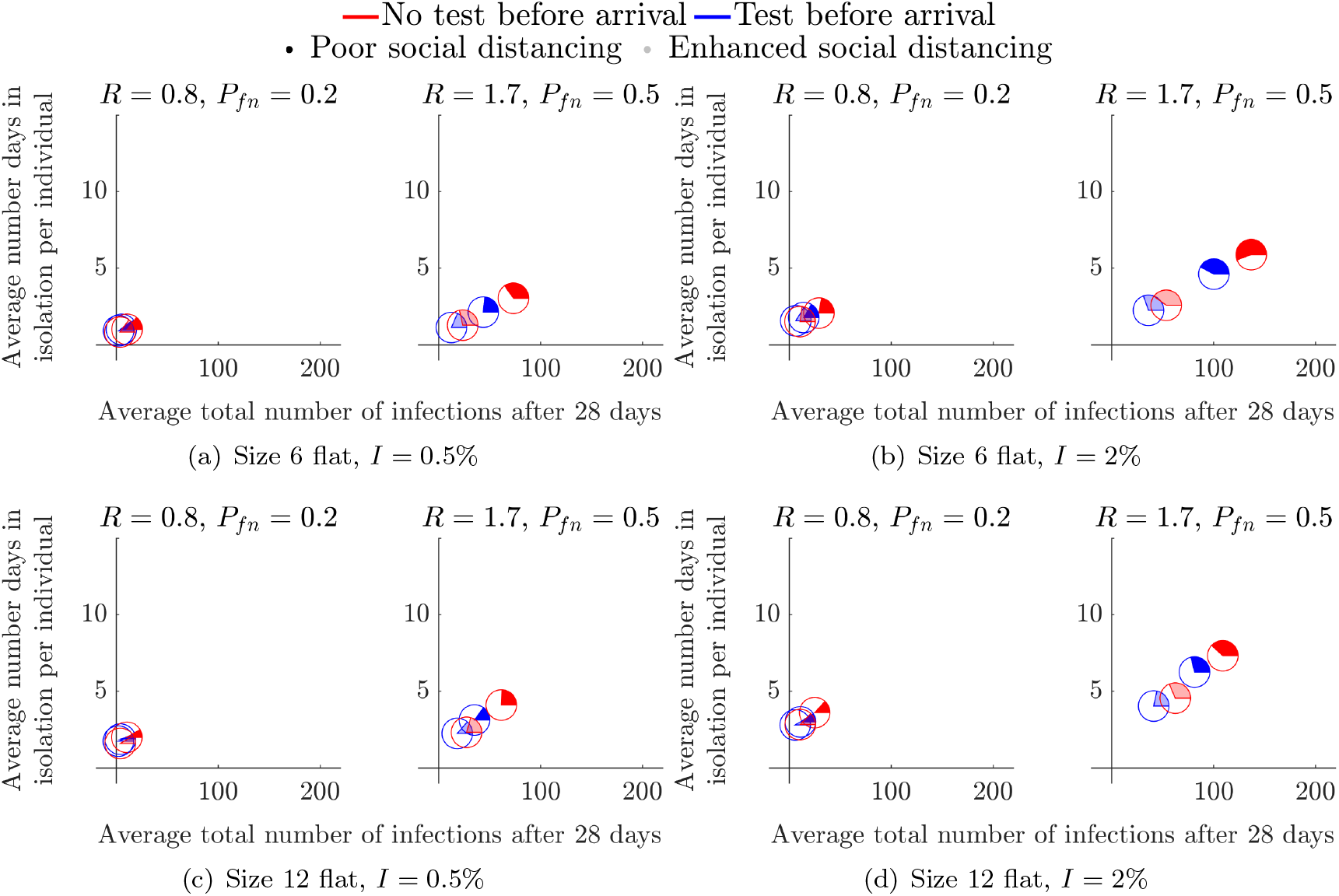
Considering the effects of a pre-term LFD test across flats of different occupancy (noted below each subfigure).

Figure 4 demonstrates that the addition of a test prior to arriving only has a significant benefit in high prevalence and high *R* number environments. The disparity in the total number of infections is particularly prominent in the cases of poor social distancing restrictions as the opaque pie charts are further apart (see figures 4(b) and 4(d)).

By comparing the results of Figure 4 across flat size, we observe that larger flat sizes result in a small decrease in average total number of infections, but a small increase in the average length of isolation period. Thus, although, a 12-person flat has an increased risk of someone bringing in an infection to the flat, once a positive test result occurs we remove more non-infected people from circulation. However, because we are only tracking infections to the point of isolation, we must be cautious regarding this interpretation. Specifically, once a flat has been isolated unless all remaining healthy individuals practice extremely good hygiene it is highly likely that most in the flat will succumb to the infection, meaning that the small reduction in total number of infections that is apparent in the results would not exist and in fact the larger flats would lead to a gain in average total of infections. Critically, this comes down to the responsibility of the individuals of an infected flat. Once a positive test has been received the flat should be clearly informed of their options and best practices that will keep the individuals safe.

In all cases we see that the coloured part of the pie chart is in the minority. Thus, at the point of isolation, we are isolating more healthy people than infected people. In particular, in larger flat sizes over 70% of all students isolating in every scenario are healthy in the 12-person flat simulations (see figures 4(c) and 4(d)). Though, it is not substantially less in the smaller 6-person flat, it is worth noting that increasing the flat size increases the number of healthy people isolating.

#### 3.2.2 Middle of term

We next consider the situation where students have returned and are continuously mixing for a 28-day period. In every infection scenario we simulated, we show that increasing the testing frequency greatly reduces the total number of infections, *i*.*e*., the green and black pie charts are the closest and furthest markers from the origin, respectively, in all subplots of Figure 5. Further, in all subplots of Figure 5, we observe the benefit of including spatial compartments to the model by the restriction of social interactions from the whole population (opaque markers) to ‘kitchen groups’ (transparent markers), particularly in the cases of *R* = 1.7, as the spatial component of the model considers the allocation of infections between the infective’s group and rest of the student population (see Section 2). Critically, when the *R* number is large, we have a distinct disparity between the opaque and transparent markers, thus, enhanced social distancing measures reduces both the number of infections and days in isolation.

**Figure 5:**
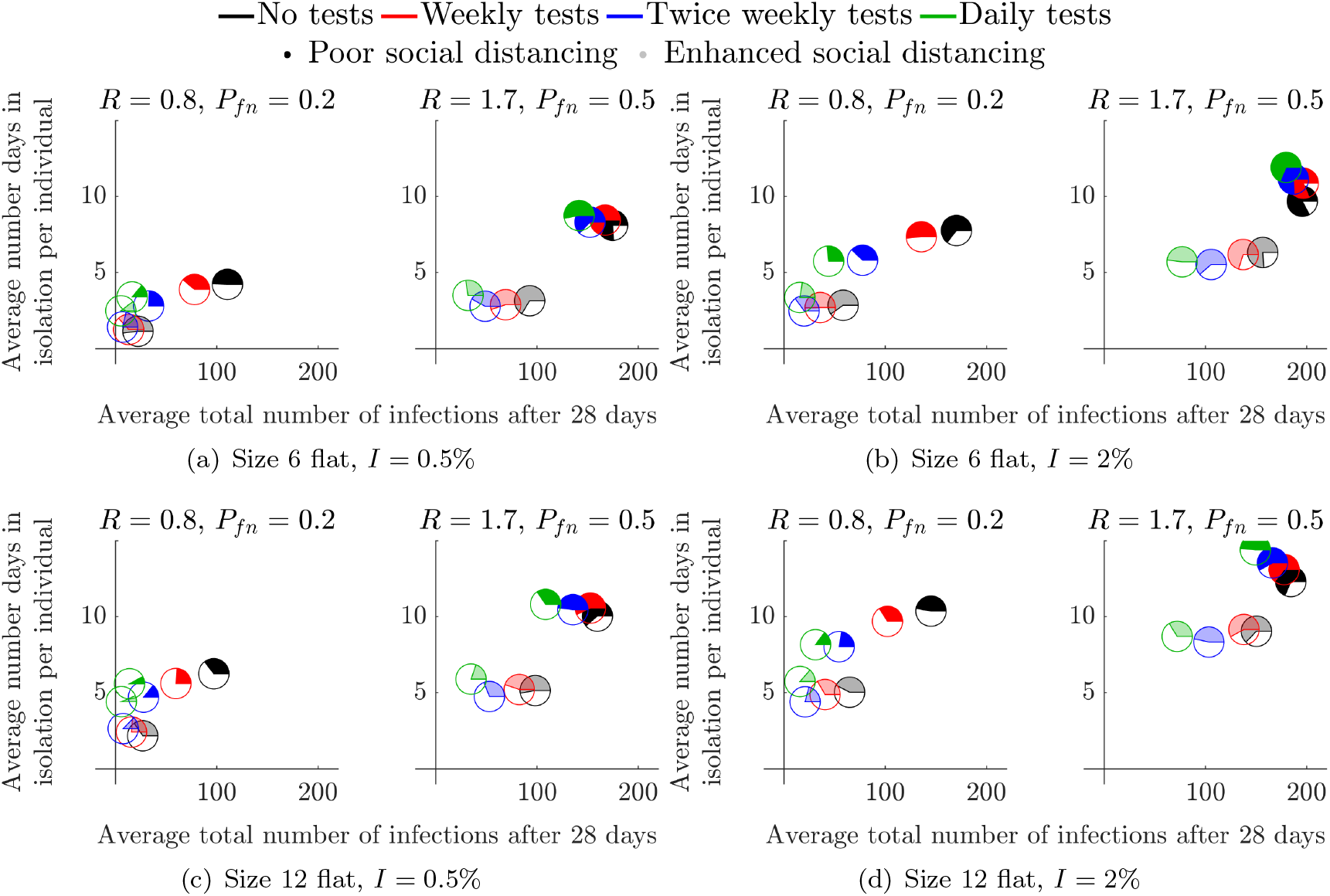
Considering the effects of a mid-term LFD test across flats of different occupancy (noted below each subfigure).

**Figure 6:**
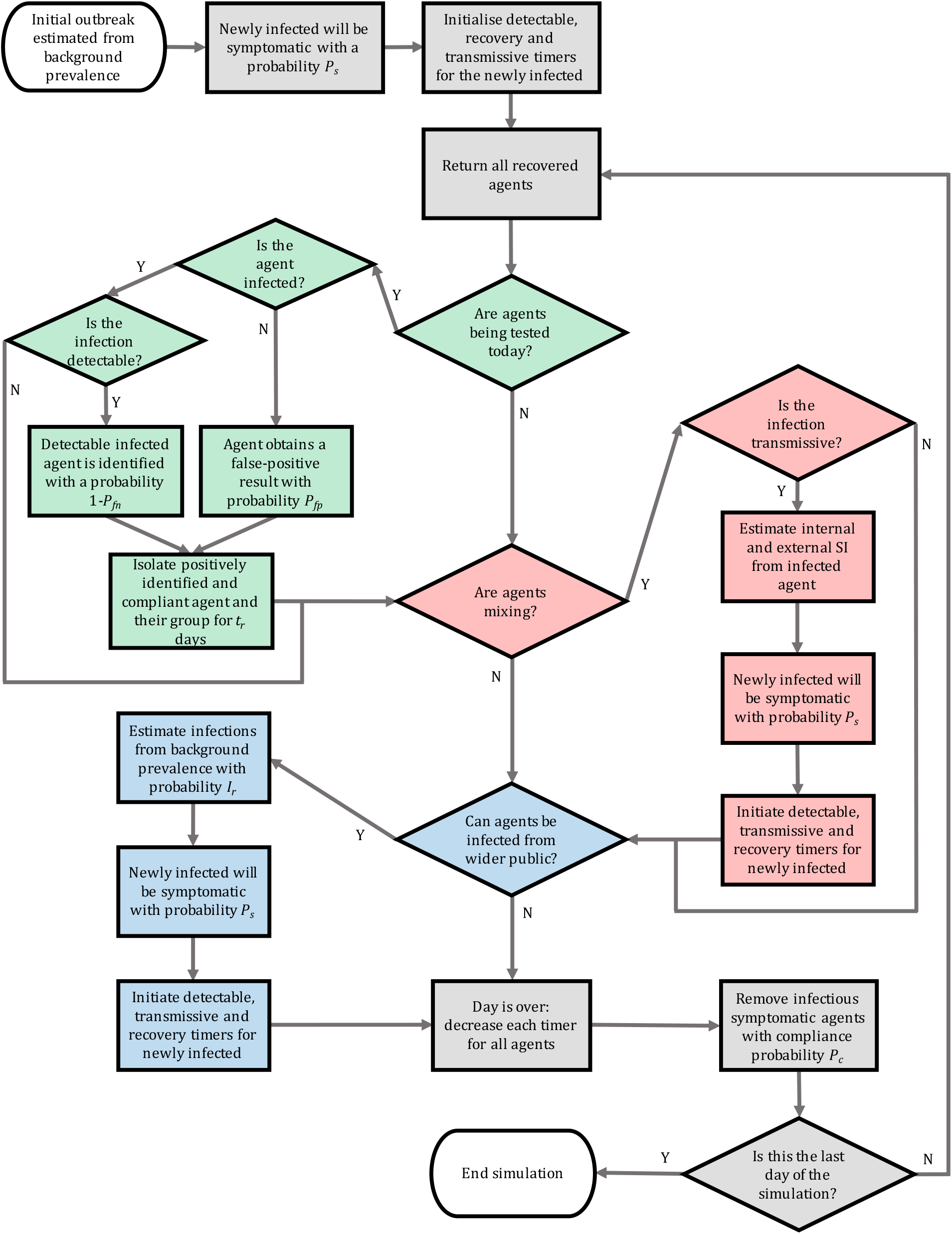
Flowchart illustrating the algorithm developed for the individual based model of infection spread. We highlight various subroutines using colours: grey is base infection routines, green is testing, red is secondary infection estimation and blue represents infections from outside the population. SI corresponds to secondary infections.

Critically, as in the secondary school case of Section 3.1, although testing is able to reduce the average total number of infected individuals it is much better to reduce the *R* value through encouraging enhanced social distancing measures. Specifically, in all cases, the transparent black pie chart is always closer to the origin than the green opaque pie chart.

Finally, our simulations suggest that increasing the flat size improves the efficacy of the enhanced social distancing measures, as all transparent pie charts are closer to the origin in the 12-person flats than their associated pie chart in the 6-person flat simulations. This observation is explained by the fact that increasing the size of a flat increases the number of healthy students that have to isolate when at least one infected student is found. Again, as mentioned in the previous section, we should critically examine this claim because unless the isolated individuals practice good hygiene the infection will spread through the isolating flat, thus generating more cases than expected. Future iterations of the model will contain this additional ability, see Section 4.2.

## 4 Discussion

Due to the time delay between becoming infectious and being detectable by a LFD [43], testing is never going to be able to stop infection spread, no matter how often it occurs. This would even be true in the case of PCR tests, which are more accurate (in terms of false negatives) and can produce results earlier in the incubation cycle. However, the time delay in obtaining results from RT-qPCR assays means that there may still be approximately one day between becoming infectious and becoming detectable. The only benefit would be a reduction in the proportion of false negatives (*P*_*fn*_), which, as we have seen, is not an important parameter to focus on.

We have observed that testing can help reduce the total number of infections as LFDs enable us to find asymptomatic individuals, who would be able to spread the disease, even if they themselves do not suffer. Although testing as much as possible will partially mitigate transmission, we must turn to alternative interventions if we want to have a significant impact on disease spread.

Here we have focused on several interventions across diverse educational contexts, both classrooms and Halls of Residence. Throughout all of our results we have seen that the simplest and cheapest way of reducing infection numbers is simply to isolate larger groups of individuals. At the extreme, isolating the entire population ensures that infections cannot be transmitted between individuals. Of course, such extreme solutions are unlikely to be desirable because we isolate many healthy individuals, whereby they miss educational and social aspects of everyday life [58]. Further, the ability to apply such a lockdown is impossible as “isolation fatigue” sets in and people’s compliance with harsh rules will likely diminish [59, 60].

Thus, how, when and who should be isolated are all important questions that should be carefully considered. Critically, isolating individual flats, classes or table groups hugely reduces the average total number of infections. However, this does not dictate total isolation, but encourages effective social distancing to limit infection spread.

We now look towards more proactive interventions that have a financial cost, namely: supplying personal protective equipment (*e*.*g*. masks); or investing in classroom sized ventilation units. Clearly, from the results of Section 3 we observe that the interventions we consider are always going to have positive influence on reducing disease transmission. Moreover, including multiple interventions offers bigger gains than each individual intervention. Based on the work of [41], we were able to show that although masks are a cheap and simple way of reducing disease spread, additional ventilation may be superior to masks in their ability to reduce the total number infections.

Specifically, additional ventilation influences our simulations by drastically reducing the localised density of contagion in the air. Though the additional air flow increases the spread of the contagion, the lower overall density greatly reduces the probability of infection for any individual both local and non-local, thus, it reduces the number of secondary infections that can occur overall. Namely, with good room ventilation test-to-release becomes a viable option (see the blue pie charts in Figure 3). Specifically, even under worst-case parameter values, good ventilation means that we only need to isolate table groups to ensure the greatest reduction in total cases.

### 4.1 Impact

Our work has already helped to influence Welsh Government policy in relation to the development of FE and HE workplace and residential policy. Our model was used to assess the impact of testing and interventions for students returning to colleges, or their Halls of Residence from their permanent home addresses. Our analysis was considered by the Welsh Government’s Technical Advisory Group (TAG), Further Education and Higher Education Task Group dealing with Covid-19. Further, the work was also presented to the Environmental Science policy committee, for use in advising how to open up more general social spaces, such as places of worship. Our work has also been communicated to colleagues in the English Government, Scottish Government and Northern Ireland Executive and is currently being used by Bethan Cradock, (Head of Policy HE Covid-19) and Marian Jebb (head of post-16 quality and data management for the Welsh Government) to develop policy for safely returning students back to their places of study.

### 4.2 Future work

Due to the continued existence of the pandemic there is still plenty be done. Much of the work could focus on making the simulation more accurate. Namely, rather than fixing parameters we could use a Bayesian approach to sample from realistic parameter distributions, which can be generated from data [61, 62, 63, 64]. Equally, current research is focused on generating estimates of real time simulations of airborne particle spread. This could be encompassed into our simulation, but we would need to increase the time resolution from days to hours.

The quickest gain for making the simulation more realistic, particularly in the HE case, would be to nest the algorithm within itself. Nesting the algorithm within itself would give us access to multiple spatial compartments. Namely, one level of the algorithm could be running a Halls of Residence on the scale of grouping everyone into flats, then a second level of the algorithm could be running on the scale of individuals within flats. This would allow the removal of the current restriction of tracking infections up until the point of group isolation.

The development of a nested algorithm would also allow us to simulate infection propagation over multiple classes within the same school. As a result, we would be able to elucidate the impact of the teachers moving from class to class, *i*.*e*. the existence of “super spreaders” from internal and external class interactions. Namely, in a next iteration we could track not only when someone is infected, but who infected them. We could then extract the number of secondary infections linked to each infected person and observe whether there are specific individuals that infect others at a rate significantly larger than others. From this point, we could reverse engineer the situations in which the super spreaders find themselves, to see if there are any commonalities which could be perturbed leading to a reduction of their highly infectious nature. Furthermore, the construction a transmission network between all agents would allow for further analysis on the existence of a “super spreader” within the population by determining connectivity bottlenecks from its spectral properties [65].

### 4.3 Summary

We asked whether it was suitable for Lateral Flow Devices (LFD) to be used as a means of getting students back to school, or university. Our simulated results show repeated testing does help reduce the average number of total infections, as asymptomatic individuals can be found and isolated, resulting in the reduction of infectious individuals. However, we have also found that it is not worth investing in better tests that reduce the false positive probability of the LFD, which has been its major criticism. Instead, time, effort and money are better spent investing in personal protective equipment (*e*.*g*. masks) and increasing the quality of ventilation in enclosed environments.

## Data Availability

No clinical data was produced in this study. We provide an open-access repository for the code and test files required to run the model outlined in the present study.

https://github.com/joshwillmoore1/COVID-19_Intervention_IBM

## 5 Acknowledgements

JWM is supported by Knowledge Economy Skills Scholarships (KESS2), a pan-Wales higher-level skills initiative led by Bangor University on behalf of the Higher Education sector in Wales. It is part-funded by the Welsh Government’s European Social Fund (ESF). JWM also received support from Just One Giant Lab (JOGL) (Grant No: 520772).

## A Individual based model interpretation and implementation

As we developed our computational framework to be used independent of technical background, we supply a list of example real-world infection events and how these events are interpreted within our model (see Table 3). We intend for the model to be analysed, adapted and extended to suit the local requirements, thus Table 3 should allow a user to prescribe the inputs required in their particular case.

Figure 6 provides a flowchart depicting the full algorithm for infection propagation throughout a discrete population, colour-coded in agreement with Figure 1 to highlight optional sub-routines of the algorithm. In addition we provide Table 4, which presents a summary of parameter values used in each scenario in Section 3. Finally, Table 5 provides the reader with definitions of every input parameter to the model.

## B Open-access online COVID-19 Intervention Simulator

An online simulator was developed to encourage the use of the stochastic agent-based model for policymakers (https://bit.ly/CV19_INTER_IBM). Namely, the applet requires no previous coding experience to operate and allows the user to reproduce the results presented in this study in addition to testing new scenarios with current infection data. The online applet contains all features of the original model which are outlined in Section 2 with some additional features to aid the decision process of policy surrounding NPIs as the circumstances evolve throughout the pandemic. The additional features of the applet are as follows:

1. Immunity of individuals in the population (vaccinations / recent infections);
2. Optional automatic Welsh infection data retrieval (included to aid the Welsh TAG);
3. Export simulation input data and output data into a downloadable excel document for further analysis.

The inclusion of these features allow the users to have up to date model predictions as new data is presented, and therefore reduce the time between data collection and policy action.

The applet was developed in collaboration with the Welsh TAG education policy sector to improve usability among policy-makers. In addition, tutorial videos are embedded into the applet to ensure appropriate usage of the software. Figure 7 demonstrates an application of the applet testing the use of masks in a large population.

**Figure 7:**
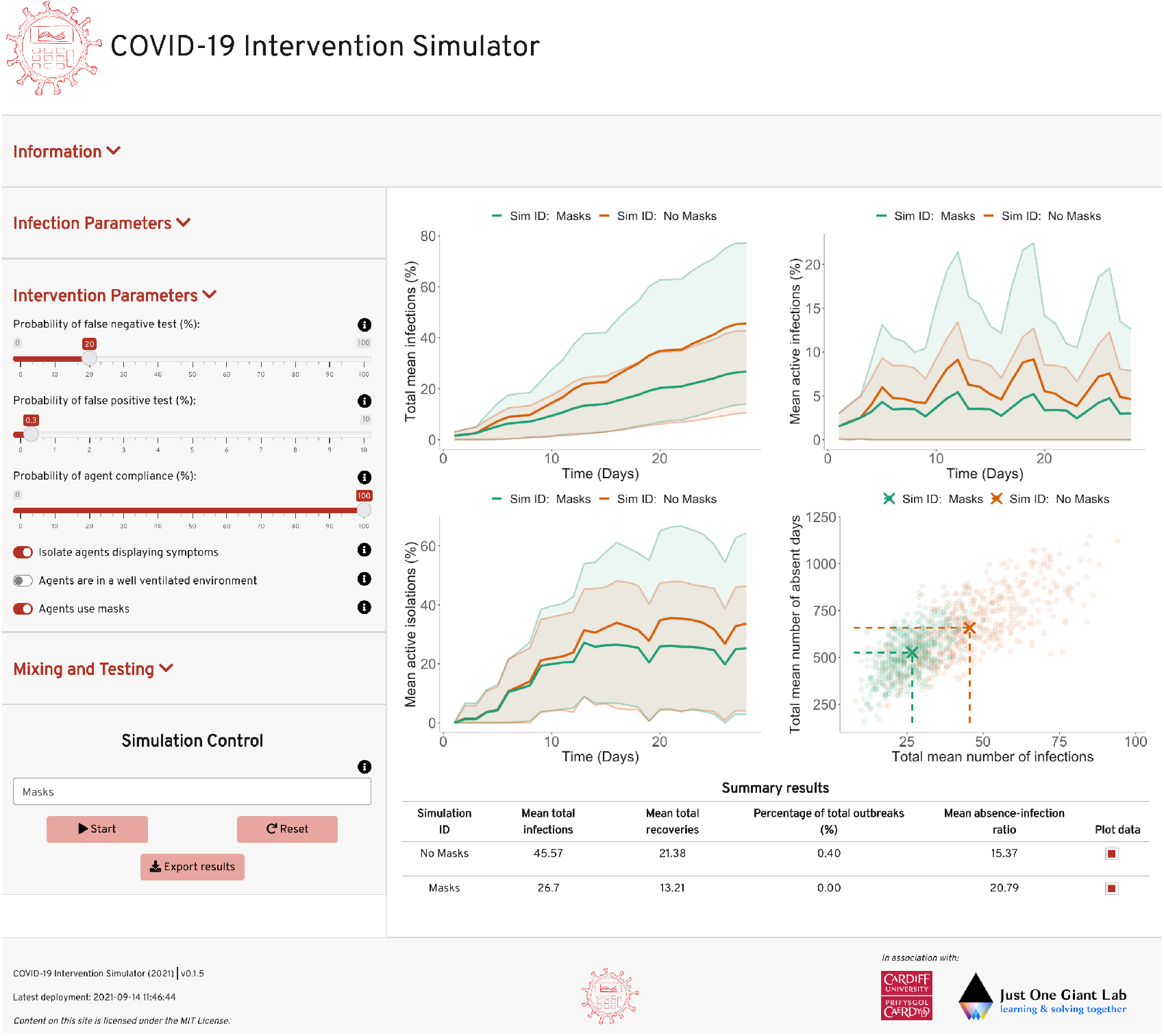
An application of the COVID-19 Intervention Simulator to compare the effect of masks on infection transmission in a population of 100 individuals that regularly mix with the wider community over the period of a month.

## C NPI parameter estimation from airborne transmission model

The NPI parameter values presented in Table 1 where estimated by simulating the semi-analytical solutions of equations 1 and 2 in a characteristic classroom environment using model parameters in Table 6 as in [41]. Specifically, an infectious person is placed at the centre of the room and the concentration of contagion particles are simulated at locations 2 (local) and 4 (nonlocal) metres away from the centre, over a period of 5 hours. These simulations were conducted with all combinations of masks and overhead ventilation (Figure 8) for both the best and worst-case infectious scenarios of Table 2. The full set of results can be found in tables 7-10. For explicit details on model construction and applications, see [41]. We note that the authors provide open-access code to simulate the airborne transmission model (equations 1 and 2) in their maintained repository at https://github.com/zechlau14/Modelling-Airborne-Transmission.

**Table 6:** Parameter estimations for running the model in [41].

| Parameter | Value | Source |
| --- | --- | --- |
| Classroom Dimensions | 8 m × 8 m × 3 m | |
| Airflow speed | 0.15 m/s | [48] |
| Room air exchange rate (ACH) | Very poor ventilation: 0.12 h<sup>-1</sup><br>Poor ventilation: 0.72 h<sup>-1</sup><br>Pre-pandemic recommended ventilation: 3 h<sup>-1</sup><br>Updated recommended ventilation: 6 h<sup>-1</sup> | [66]<br>[66]<br>[67]<br>[68] |
| Eddy diffusion coefficient | Very poor ventilation: 8.8 $\times 10^{-4}$ m<sup>2</sup>/s<br>Poor ventilation: 5.3 $\times 10^{-3}$ m<sup>2</sup>/s<br>Pre-pandemic recommended ventilation: 2.2 $\times 10^{-2}$ m<sup>2</sup>/s<br>Updated recommended ventilation: 4.4 $\times 10^{-2}$ m<sup>2</sup>/s | [50] |
| Breathing rate | 1.3 $\times 10^{-4}$ m<sup>3</sup>/s | [69] |
| Generation rate of infectious particles | At rest: 0.5 particles/s<br>Low activity: 1.4 particles/s<br>High activity: 5 particles/s | [70] |
| Efficiency of mask | 0.5 | [49] |
| Virus deactivation rate | 1.7 $\times 10^{-4}$ s<sup>-1</sup> | [71] |
| Aerosol settling rate | 1.1 $\times 10^{-4}$ s<sup>-1</sup> | [72] |
| Median infectious dose | 100 particles | [73] |

**Table 7:** Steady state local and non-local concentration of airborne contagions in a secondary school classroom with varying ventilation levels. Calculated from the model in [41] and the parameters in Table 6.

| Activity | ACH = 0.12 |  | ACH = 0.72 |  | ACH = 3.00 |  | ACH = 6.00 |  |
| --- | --- | --- | --- | --- | --- | --- | --- | --- |
| | $A_l$ | $A_n$ | $A_l$ | $A_n$ | $A_l$ | $A_n$ | $A_l$ | $A_n$ |
| At rest with mask | 21.1 | 3.7 | 8.2 | 4.3 | 3.1 | 2.0 | 1.7 | 1.2 |
| At rest no mask | 42.2 | 7.3 | 16.4 | 8.6 | 6.2 | 4.1 | 3.5 | 2.3 |
| Low activity with mask | 59.4 | 10.2 | 23.0 | 12.0 | 8.7 | 5.8 | 4.8 | 3.3 |
| Low activity no mask | 118.3 | 20.5 | 46.0 | 24.1 | 17.5 | 11.5 | 9.7 | 6.6 |
| High activity / Superspreader with mask | 202.8 | 35.1 | 78.8 | 41.3 | 30 | 19.8 | 16.6 | 11.2 |
| High activity / Superspreader no mask | 422.4 | 73.1 | 164.2 | 86.1 | 62.5 | 41.2 | 34.6 | 23.4 |

**Table 8:** Local and nonlocal infection risk from airborne transmission after 5 hours in a secondary school classroom with varying ventilation levels. Calculated from the model in [41] and the parameters in Table 6.

| Activity | ACH = 0.12 |  | ACH = 0.72 |  | ACH = 3.00 |  | ACH = 6.00 |  |
| --- | --- | --- | --- | --- | --- | --- | --- | --- |
| | $I_{Al}$ | $I_{An}$ | $I_{Al}$ | $I_{An}$ | $I_{Al}$ | $I_{An}$ | $I_{Al}$ | $I_{An}$ |
| At rest with mask | 0.267 | 0.041 | 0.115 | 0.058 | 0.047 | 0.031 | 0.027 | 0.018 |
| At rest no mask | 0.462 | 0.080 | 0.217 | 0.113 | 0.092 | 0.061 | 0.053 | 0.036 |
| Low activity with mask | 0.581 | 0.110 | 0.289 | 0.154 | 0.127 | 0.084 | 0.074 | 0.050 |
| Low activity no mask | 0.824 | 0.207 | 0.495 | 0.284 | 0.238 | 0.161 | 0.142 | 0.097 |
| High activity / Superspreader with mask | 0.949 | 0.329 | 0.690 | 0.436 | 0.372 | 0.260 | 0.231 | 0.161 |
| High activity / Superspreader no mask | 0.998 | 0.534 | 0.913 | 0.697 | 0.621 | 0.466 | 0.421 | 0.306 |

**Table 9:** Worst-case local *R* number, *R*_*l*_ and nonlocal *R* number, *R*_*n*_ from airborne transmission in a secondary school classroom with varying ventilation levels. Calculated from the relation *R*_*l*_ = *I*_*Al*_*R/*(*I*_*Al*_+ *IAn*) and *R*_*n*_ = *I*_*An*_*R/*(*I*_*Al*_ + *IAn*), and the values in Table 8 and the worst-case scenario *R*-value given in Table 2.

| Activity | ACH = 0.12 |  | ACH = 0.72 |  | ACH = 3.00 |  | ACH = 6.00 |  |
| --- | --- | --- | --- | --- | --- | --- | --- | --- |
| | $R_l$ | $R_n$ | $R_l$ | $R_n$ | $R_l$ | $R_n$ | $R_l$ | $R_n$ |
| At rest with mask | 1.47 | 0.23 | 1.13 | 0.57 | 1.02 | 0.68 | 1.02 | 0.68 |
| At rest no mask | 1.45 | 0.25 | 1.12 | 0.58 | 1.02 | 0.68 | 1.01 | 0.69 |
| Low activity with mask | 1.43 | 0.27 | 1.11 | 0.59 | 1.0 | 0.68 | 1.01 | 0.69 |
| Low activity no mask | 1.36 | 0.34 | 1.08 | 0.62 | 1.01 | 0.69 | 1.01 | 0.69 |
| High activity / Superspreader with mask | 1.26 | 0.44 | 1.04 | 0.66 | 1.00 | 0.70 | 1.00 | 0.70 |
| High activity / Superspreader no mask | 1.11 | 0.59 | 0.96 | 0.74 | 0.97 | 0.73 | 0.98 | 0.72 |

**Table 10:** Best-case local *R* number, *R*_*l*_ and nonlocal *R* number, *R*_*n*_ from airborne transmission in a secondary school classroom with varying ventilation levels. Calculated from the relation *R*_*l*_ = *I*_*Al*_*R/*(*I*_*Al*_+ *IAn*) and *R*_*n*_ = *I*_*An*_*R/*(*I*_*Al*_ + *IAn*), and the values in Table 8 and the best-case scenario *R*-value given in Table 2.

| Activity | ACH = 0.12 |  | ACH = 0.72 |  | ACH = 3.00 |  | ACH = 6.00 |  |
| --- | --- | --- | --- | --- | --- | --- | --- | --- |
| | $R_l$ | $R_n$ | $R_l$ | $R_n$ | $R_l$ | $R_n$ | $R_l$ | $R_n$ |
| At rest with mask | 0.69 | 0.11 | 0.53 | 0.17 | 0.48 | 0.32 | 0.48 | 0.32 |
| At rest no mask | 0.68 | 0.12 | 0.53 | 0.27 | 0.48 | 0.32 | 0.48 | 0.32 |
| Low activity with mask | 0.67 | 0.13 | 0.52 | 0.28 | 0.48 | 0.32 | 0.48 | 0.32 |
| Low activity no mask | 0.64 | 0.16 | 0.51 | 0.29 | 0.48 | 0.32 | 0.48 | 0.32 |
| High activity / Superspreader with mask | 0.59 | 0.21 | 0.49 | 0.1 | 0.47 | 0.33 | 0.47 | 0.33 |
| High activity / Superspreader no mask | 0.52 | 0.28 | 0.45 | 0.35 | 0.46 | 0.34 | 0.46 | 0.34 |

**Figure 8:**
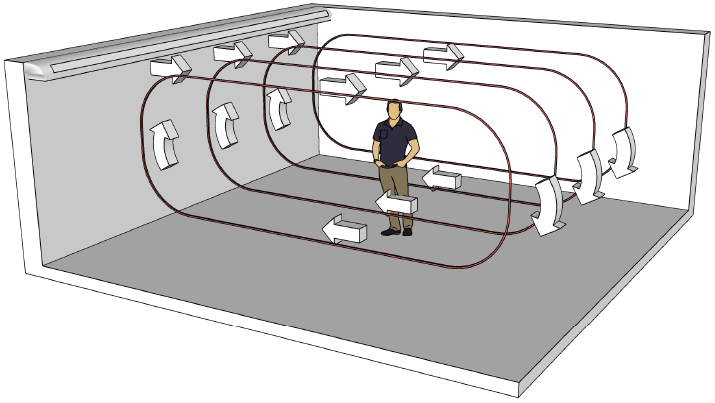
Recirculating airflow in the simulated classroom.

## Footnotes

1 Simulations were run on a 2.6 GHz 6-Core Intel Core i7 with 16 GB 2667 MHz DDR4 2019 MacBook Pro.

## References

[1] C. Wang, P. W. Horby, F. G. Hayden, and G. F. Gao. A novel coronavirus outbreak of global health concern. The Lancet, 395(10223):470–473, 2020.

[2] M. Chahrour, S. Assi, M. Bejjani, A. A. Nasrallah, H. Salhab, M. Fares, and H. H. Khachfe. A bibliometric analysis of covid-19 research activity: A call for increased output. Cureus, 12(3), 2020.

[3] P. Sahu. Closure of universities due to coronavirus disease 2019 (covid-19): impact on education and mental health of students and academic staff. Cureus, 12(4), 2020.

[4] C. Rapanta, L. Botturi, P. Goodyear, L Guàrdia, and M. Koole. Online university teaching during and after the covid-19 crisis: Refocusing teacher presence and learning activity. Postdigi. Sci. Edu., pages 1–23, 2020.

[5] Education, universities and childcare.. URL https://www.gov.uk/coronavirus/education-and-childcare.

[6] Schools: coronavirus guidance.. URL https://gov.wales/schools-coronavirus-guidance.

[7] Higher education: coronavirus.. URL https://gov.wales/schools-coronavirus-guidance.

[8] Spi-m-o: Summary of modelling on scenarios for easing restrictions,. URL https://assets.publishing.service.gov.uk/government/uploads/system/uploads/attachmentdata/file/963400/S1116_SPI-M-O_Summary_of_modelling_on_scenario_for_easing_restrictions.pdf.

[9] H. Melnick and L. Darling-Hammond. Reopening schools in the context of covid-19: Health and safety guidelines from other countries. policy brief. Learning Policy Institute, 2020.

[10] J. M. Donohue and E. Miller. Covid-19 and school closures. JAMA, 324(9):845–847, 2020.

[11] J. Daniel. Education and the covid-19 pandemic. Prospects, 49(1):91–96, 2020.

[12] I. Asanov, F. Flores, D. McKenzie, M. Mensmann, and M. Schulte. Remote-learning, time-use, and mental health of ecuadorian high-school students during the covid-19 quarantine. World development, 138:105225, 2021.

[13] E. Amodio, M. Battisti, A. Kourtellos, G. Maggio, and C. M. Maida. Schools opening and covid-19 diffusion: Evidence from geolocalized microdata1. COVID Econom.

[14] Covid-19 in children and the role of school settings in transmission. URL https://www.ecdc.europa.eu/en/publications-data/children-and-school-settings-covid-19-transmission.

[15] R. Lordan, G. A. FitzGerald, and T. Grosser. Reopening schools during covid-19. Science, 369 (6508):1146–1146, 2020. ISSN 0036-8075. doi: 10.1126/science.abe5765. URL https://science.sciencemag.org/content/369/6508/1146.

[16] Lateral flow testing in schools and colleges faqs. URL https://www.ascl.org.uk/Help-and-Advice/Leadership-and-governance/Health,-safety-and-safeguarding/Coronavirus-essential-information/Coronavirus-FAQs/Lateral-flow-testing-in-schools-and-colleges.

[17] Coronavirus: testing in schools and colleges. URL https://neu.org.uk/coronavirus-school-testing.

[18] Routine testing for education and childcare staff.. URL https://gov.wales/routine-testing-education-and-childcare-staff-html.

[19] I. Torjesen. Covid-19: How the uk is using lateral flow tests in the pandemic. BMJ, 372, 2021. doi: 10.1136/bmj.n287. URL https://www.bmj.com/content/372/bmj.n287.

[20] Coronavirus (covid-19): Test to release for international travel.. URL https://www.gov.uk/guidance/coronavirus-covid-19-test-to-release-for-international-travel#how-the-scheme-works.

[21] J. Deeks, M. Gill, S. Bird, S. Richardson, and D. Ashby. Covid-19 INNOVA testing in schools: don’t just test, evaluate, January 2021. URL https://blogs.bmj.com/bmj/2021/01/12/covid-19-innova-testing-in-schools-dont-just-test-evaluate/.

[22] Preliminary report from the joint phe porton down & university of oxford sars-cov-2 test development and validation cell: Rapid evaluation of lateral flow viral antigen detection devices (lfds) for mass community testing:. URL https://www.ox.ac.uk/sites/files/oxford/media_wysiwyg/UKevaluation_PHEPortonDownUniversityofOxford_final.pdf.

[23] Scientific Advisory Group for Emergencies. SPI-B: Possible impact of the COVID-19 vaccination programme on adherence to rules and guidance about personal protective behaviours aimed at preventing spread of the virus, 17 December 2020, 2020. URL https://www.gov.uk/government/publications/spi-b-possible-impact-of-the-covid-19-vaccination-programme-on-adherence-to-rules-and-guidance-about-personal-protective-behaviours-aimed-at-preventi.

[24] Moriah Bergwerk, Tal Gonen, Yaniv Lustig, Sharon Amit, Marc Lipsitch, Carmit Cohen, Michal Mandelboim, Einav Gal Levin, Carmit Rubin, Victoria Indenbaum, et al. Covid-19 breakthrough infections in vaccinated health care workers. New England Journal of Medicine, 2021.

[25] Fiona P Havers, Huong Pham, Christopher A Taylor, Michael Whitaker, Kadam Patel, Onika Anglin, Anita K Kambhampati, Jennifer Milucky, Elizabeth Zell, Shua J Chai, et al. Covid-19-associated hospitalizations among vaccinated and unvaccinated adults≥ 18 years–covid-net, 13 states, january 1–july 24, 2021. medRxiv, 2021.

[26] Centers fro Disease Control and Prevention.Your guide to masks, 2021. URL https://www.cdc.gov/coronavirus/2019-ncov/prevent-getting-sick/about-face-coverings.html.

[27] Bernadette C Young, David W Eyre, Saroj Kendrick, Chris White, Sylvester Smith, George Beveridge, Toby Nonnemacher, Fegor Ichofu, Joseph Hillier, Ian Diamond, et al. A cluster randomised trial of the impact of a policy of daily testing for contacts of covid-19 cases on attendance and covid-19 transmission in english secondary schools and colleges. medRxiv, 2021.

[28] ONS. Coronavirus (covid-19) latest insights: Infections, 2021. URL https://www.ons.gov.uk/peoplepopulationandcommunity/healthandsocialcare/conditionsanddiseases/articles/coronaviruscovid19latestinsights/infections.

[29] UK Health Security Agency. The latest reproduction number (r) and growth rate of coronavirus (covid-19), 2021. URL https://www.gov.uk/guidance/the-r-value-and-growth-rate.

[30] Nicholas G Davies, Petra Klepac, Yang Liu, Kiesha Prem, Mark Jit, and Rosalind M Eggo. Age-dependent effects in the transmission and control of covid-19 epidemics. Nature medicine, 26(8): 1205–1211, 2020.

[31] C. S. M. Currie, J. W. Fowler, K. Kotiadis, T. Monks, B. S. Onggo, D. A. R., and A. A. Tako. How simulation modelling can help reduce the impact of covid-19. Journal of Simulation, pages 1–15, 2020.

[32] N. M. Ferguson, D. Laydon, G. Nedjati-Gilani, N. Imai, K. Ainslie, M. Baguelin, S. Bhatia, A. Boonyasiri, Z. Cucunubá, G. Cuomo-Dannenburg, and A. Dighe. Impact of non-pharmaceutical interven-tions (npis) to reduce covid-19 mortality and healthcare demand. Imperial College COVID-19 Response Team. URL https://www.imperial.ac.uk/media/imperial-college/medicine/sph/ide/gida-fellowships/Imperial-College-COVID19-NPI-modelling-16-03-2020.pdf.

[33] S. Latif, M. Usman, S. Manzoor, W. Iqbal, J. Qadir, G. Tyson, I. Castro, A. Razi, M. N. K. Boulos, and A. Weller. Leveraging data science to combat covid-19: A comprehensive review. 2020.

[34] P. R. Harper, J. W. Moore, and T. E. Woolley. Covid-19 transmission modelling of students returning home from university. Health Systems, 0(0):1–10, 2021. doi: 10.1080/20476965.2020.1857214. URL https://doi.org/10.1080/20476965.2020.1857214.

[35] A. Zhigljavsky, R. Whitaker, I. Fesenko, K. Kremnizer, J. Noonan, P. Harper, J. Gillard, T. Woolley, D. Gartner, and J. Grimsley. Generic probabilistic modelling and non-homogeneity issues for the uk epidemic of covid-19. arXiv preprint 2004.01991, 2020.

[36] P. Auger and A. Moussaoui. On the threshold of release of confinement in an epidemic seir model taking into account the protective effect of mask. Bulletin of mathematical biology, 83(4):1–18, 2021.

[37] SPI-M modelling summary for pandemic influenza,. URL https://www.gov.uk/government/publications/spi-m-publish-updated-modelling-summary.

[38] Matt J Keeling, Michael J Tildesley, Benjamin D Atkins, Bridget Penman, Emma Southall, Glen Guyver-Fletcher, Alex Holmes, Hector McKimm, Erin E Gorsich, Edward M Hill, et al. The impact of school reopening on the spread of covid-19 in england. medRxiv, 2020.

[39] C. Sun and Z. Zhai. The efficacy of social distance and ventilation effectiveness in preventing covid-19 transmission. Sust. Citi. Soc., 62:102390, 2020.

[40] UK Government. Covid-19 in the uk, 2021. URL https://coronavirus.data.gov.uk/details/download.

[41] Z. Lau, I.M. Griffiths, A. English, and K. Kaouri. Predicting the spatially varying infection risk in indoor spaces using an efficient airborne transmission model. 2021. URL https://arxiv.org/abs/2012.12267.

[42] ONS. Coronavirus (covid-19) infection survey, uk: 19 february 2021. accessed 24th February. URL https://www.ons.gov.uk/peoplepopulationandcommunity/healthandsocialcare/conditionsanddiseases/bulletins/coronaviruscovid19infectionsurveypilot/19february2021/pdf.

[43] A. Crozier, S. Rajan, I. Buchan, and M. McKee. Put to the test: use of rapid testing technologies for covid-19. BMJ, 372, 2021.

[44] J. J. Deeks and A. E. Raffle. Lateral flow tests cannot rule out sars-cov-2 infection. BMJ, 371, 2020. doi: 10.1136/bmj.m4787. URL https://www.bmj.com/content/371/bmj.m4787.

[45] Y. Cheng, N. Ma, C. Witt, S. Rapp, P.S. Wild, M.O. Andreae, U. Pöschl, and H. Su. Face masks effectively limit the probability of SARS-CoV-2 transmission. Science, 2021.

[46] G Correia, L Rodrigues, M Gameiro Da Silva, and T Gonçalves. Airborne route and bad use of ventilation systems as non-negligible factors in SARS-CoV-2 transmission. Medical hypotheses, 141: 109781, 2020.

[47] B. Zhao, Y. Liu, and C. Chen. Air purifiers: A supplementary measure to remove airborne SARS-CoV-2. Building and Environment, 177:106918, 2020.

[48] ASHRAE Standard. Standard 55-2010, thermal environmental conditions for human occupancy, 2010.

[49] E.P. Fischer, M.C. Fischer, D. Grass, I. Henrion, W.S. Warren, and E. Westman. Low-cost measurement of face mask efficacy for filtering expelled droplets during speech. Science Advances, 6 (36), 2020.

[50] T. Foat, J. Drodge, J. Nally, and S. Parker. A relationship for the diffusion coefficient in eddy diffusion based indoor dispersion modelling. Building and Environment, 169:106591, 2020.

[51] Martin Z Bazant and John WM Bush. A guideline to limit indoor airborne transmission of covid-19. Proceedings of the National Academy of Sciences, 118(17), 2021.

[52] Ehsan S Mousavi, Krystal J Godri Pollitt, Jodi Sherman, and Richard A Martinello. Performance analysis of portable hepa filters and temporary plastic anterooms on the spread of surrogate coronavirus. Building and environment, 183:107186, 2020.

[53] NHS England. Covid-19 vaccinations, 2021. URL https://www.england.nhs.uk/statistics/statistical-work-areas/covid-19-vaccinations/.

[54] Department of Health, Social Care, and Scientific Advisory Group for Emergencies. The r value and growth rate in the uk, 2020. URL https://www.gov.uk/guidance/the-r-number-in-the-uk.

[55] Z. Hyde. Covid-19, children, and schools: overlooked and at risk. Med J Aust, 213(10):444–446, 2020.

[56] H. Nishiura, T. Kobayashi, T. Miyama, A. Suzuki, S.-M Jung, K. Hayashi, R. Kinoshita, Y. Yang, B. Yuan, and A. R. Akhmetzhanov. Estimation of the asymptomatic ratio of novel coronavirus infections (covid-19). J. Infect. Dis., 94:154, 2020.

[57] M. Day. Covid-19: four fifths of cases are asymptomatic, china figures indicate. BMJ, 369, 2020. doi: 10.1136/bmj.m1375. URL https://www.bmj.com/content/369/bmj.m1375.

[58] A. Brodeur, A. E. Clark, S. Fleche, and N. Powdthavee. Covid-19, lockdowns and well-being: Evidence from google trends. Journal of public economics, 193:104346, 2021.

[59] L. J. Labrague and C. A. Ballad. Lockdown fatigue among college students during the covid-19 pandemic: Predictive role of personal resilience, coping behaviours, and health. medRxiv, 2020. doi: 10.1101/2020.10.18.20213942. URL https://www.medrxiv.org/content/early/2020/10/20/2020.10.18.20213942.

[60] S. Brouard, P. Vasilopoulos, and M. Becher. Sociodemographic and psychological correlates of compliance with the covid-19 public health measures in france. Canadian Journal of Political Science/Revue canadienne de science politique, 53(2):253–258, 2020.

[61] J. Wu, Y. Huang, C. Tu, C. Bi, Z. Chen, L. Luo, M. Huang, M. Chen, C. Tan, Z. Wang, K. Wang, Y. Liang, J. Huang, X. Zheng, and J. Liu. Household transmission of sars-cov-2, zhuhai, china, 2020. Clin. Infect. Dis., 05 2020. ISSN 1058-4838. doi: 10.1093/cid/ciaa557. URL https://doi.org/10.1093/cid/ciaa557.ciaa557.

[62] E. S. Rosenberg, E. M. Dufort, D. S. Blog, E. W. Hall, D. Hoefer, B. P. Backenson, A. T. Muse, J. N. Kirkwood, K. St. George, D. R. Holtgrave, B. J. Hutton, and H. A. Zucker. COVID-19 Testing, Epidemic Features, Hospital Outcomes, and Household Prevalence, New York State—March 2020. Clin. Infect. Dis., 71(8):1953–1959, 05 2020. ISSN 1058-4838. doi: 10.1093/cid/ciaa549. URL https://doi.org/10.1093/cid/ciaa549.

[63] N. M. Lewis, V. T. Chu, D. Ye, E. E. Conners, R. Gharpure, R. L. Laws, H. E. Reses, B. D. Freeman, M. Fajans, E. M. Rabold, P. Dawson, S. Buono, S. Yin, D. Owusu, A. Wadhwa, M. Pomeroy, A. Yousaf, E. Pevzner, H. Njuguna, K. A. Battey, C. H. Tran, V. L. Fields, P. Salvatore, M. O’Hegarty, J. Vuong, R. Chancey, C. Gregory, M. Banks, J. R. Rispens, E. Dietrich,P. Marcenac, A. M. Matanock, L. Duca, A. Binder, G. Fox, S. Lester, L. Mills, S. I. Gerber, J. Watson, A. Schumacher, L. Pawloski, N. J. Thornburg, A. J. Hall, T. Kiphibane, S. Willardson, K. Christensen, L. Page, S. Bhattacharyya, T. Dasu, A. Christiansen, I. W. Pray, R. P. Westergaard, A. C. Dunn, J. E. Tate, S. A. Nabity, and H. L. Kirking. Household Transmission of SARS-CoV-2 in the United States. Clin. Infect. Dis., 08 2020. ISSN 1058-4838. doi: 10.1093/cid/ciaa1166. URL https://doi.org/10.1093/cid/ciaa1166.ciaa1166.

[64] J. Lopez Bernal, N. Panagiotopoulos, C. Byers, T. Garcia Vilaplana, N. L. Boddington, X. Zhang, A. Charlett, S. Elgohari, L. Coughlan, R. Whillock, S. Logan, H. Bolt, M. Sinnathamby, L. Letley, P. MacDonald, R. Vivancos, O. Edeghere, C. Anderson, K. Paranthaman, S. Cottrell, J. McMenamin, M. Zambon, G. Dabrera, M. Ramsay, and V. Saliba. Transmission dynamics of covid-19 in household and community settings in the united kingdom. medRxiv, 2020. doi: 10.1101/2020.08.19.20177188. URL https://www.medrxiv.org/content/early/2020/08/22/2020.08.19.20177188.

[65] F. Chung and F. Graham. Spectral graph theory. Number 92. American Mathematical Soc., 1997.

[66] H. Guo, L. Morawska, C. He, and D. Gilbert. Impact of ventilation scenario on air exchange rates and on indoor particle number concentrations in an air-conditioned classroom. Atmospheric Environment, 42(4):757–768, 2008.

[67] ASHRAE Standard. Standard 62-2007 ventilation for acceptable indoor air quality, 2007.

[68] ASHRAE. Reopening of schools and universities. Available at: https://www.ashrae.org/technical-resources/reopening-of-schools-and-universities, 2020. [Accessed: 29 October 2020].

[69] S. Hallett, F. Toro, and J.V. Ashurst. Physiology, tidal volume, 2020. [Updated 2020 Jun 1]. In: StatPearls [Internet]. Treasure Island (FL): StatPearls Publishing; 2020 Jan-. Available from: https://www.ncbi.nlm.nih.gov/books/NBK482502/.

[70] S. Asadi, A.S. Wexler, C.D. Cappa, S. Barreda, N.M. Bouvier, and W.D. Ristenpart. Aerosol emission and superemission during human speech increase with voice loudness. Scientific Reports, 9(1):1–10, 2019.

[71] N. van Doremalen, T. Bushmaker, D.H. Morris, M.G. Holbrook, A. Gamble, B.N. Williamson, A. Tamin, J.L. Harcourt, N.J. Thornburg, S.I. Gerber, J.O. Lloyd-Smith, E. de Wit, and V.J. Munster. Aerosol and surface stability of SARS-CoV-2 as compared with SARS-CoV-1. New England Journal of Medicine, 382(16):1564–1567, 2020.

[72] P.M. de Oliveira, L.C.C. Mesquita, S. Gkantonas, A. Giusti, and E. Mastorakos. Evolution of spray and aerosol from respiratory releases: theoretical estimates for insights on viral transmission. Proc. R. Soc. A, 477:20200584, 2021.

[73] V. Vuorinen, M. Aarnio, M. Alava, V. Alopaeus, N. Atanasova, M. Auvinen, N. Balasubramanian, H. Bordbar, P. Erästö, R. Grande, et al. Modelling aerosol transport and virus exposure with numerical simulations in relation to SARS-CoV-2 transmission by inhalation indoors. Safety Science, 130:104866, 2020.

